# Fasting, ketogenic and anti-inflammatory diets stabilized active relapsing-remitting multiple sclerosis over 18 months – a randomized, controlled study

**DOI:** 10.1101/2024.08.13.24311863

**Authors:** Lina S. Bahr, Judith Bellmann-Strobl, Daniela A. Koppold, Rebekka Rust, Tanja Schmitz-Hübsch, Maja Olszewska, Jean Stadlbauer, Markus Bock, Michael Scheel, Claudia Chien, Jan Multmeier, Alexander Krannich, Andreas Michalsen, Friedemann Paul, Anja Mähler

## Abstract

**Background:** Multiple sclerosis (MS) is the most common inflammatory disease of the central nervous system in young adulthood leading to disability and early retirement. Ketone-based diets improve the disease course in MS animal models and health outcomes in different pilot studies of neurodegenerative diseases.

**Methods:** We enrolled 105 individuals with relapsing-remitting MS (RRMS) in an 18-month, randomized, controlled study, and randomized them into 1) standard healthy diet (SD) as recommended by the German Nutrition Society, 2) fasting diet (FD) with 7-day fasts every 6 months with intermittent fasting at 6 of 7 days a week or 3) ketogenic diet (KD) with 20-40 g carbohydrates per day. Primary outcome was the number of new MRI lesions after 18 months in the KD and FD compared to SD and compared to baseline. Secondary outcomes included further MRI outcomes, disease biomarkers as well as metabolic, and clinical MS outcomes.

**Results:** Eighty-one participants completed the study. The primary endpoint number of new T2 lesions after 18 months did not change in any of the groups (SD 0 (0-(−1)), FD 0 (2-0), KD 0 (2-0)). Compared to baseline, in the FD group, Neurofilament light chain (NfL) - concentrations were lower at 9 months (−1.94 pg/mL, p = 0.042) and depressive symptoms improved slightly at 18 months (p = 0.079). In the KD group, cognition improved at 18 months (symbol digit modalities test +3.7, p = 0.020). Cardiometabolic risk markers (body mass index, abdominal fat, blood lipids, adipokines, blood pressure) improved in all three groups at 9 months differently and were partially associated with clinical outcomes in the FD and KD group.

**Conclusion:** Dietary interventions may stabilize RRMS disease course and improve cardiometabolic risk factors, cognition, and depressive symptoms, providing valuable complementary treatment options.

**Trial registration:** ClinicalTrials.gov, NCT03508414. Retrospectively registered on 25 April 2018.

## Introduction

Multiple sclerosis (MS) is an autoimmune inflammatory and neurodegenerative disorder of the central nervous system caused by a combination of genetic susceptibility and environmental factors (1). Many individuals with MS are highly motivated to incorporate complementary approaches, such as specific diets, into their treatment. However, despite 70 years of research efforts in this field, there is still no evidence-based diet showing a beneficial effect on disease progression. Current dietary recommendations, such as the Mediterranean diet, aim at decreasing the risk of comorbidities, and thereby secondarily ameliorate prognosis (2). However, robust evidence of effects of healthy, anti-inflammatory diets on MS disease pathology itself is still lacking (3, 4).

In contrast, emerging data on fasting diets (FD) and ketogenic diets (KD) indicate direct effects. Both FD and KD drastically reduce carbohydrate intake, initiating ketone body production from internal and/or external fats, the state of ketosis. These ketone bodies may provide an alternative, more efficient, energy source for the brain, which might be important for the regeneration of demyelinated axons (5).

First indicators on the effectiveness of FDs in MS come from animal experimental studies. A calorie restriction of 40 percent ameliorated the clinical disease course of murine experimental autoimmune encephalomyelitis (EAE). Calorie restricted mice showed less severe inflammation, demyelination, and axonal injury (6). In addition, alternate day fasting in EAE mice reduced incidence, onset, and severity of the disease (7). A similar study confirmed these results and suggested that intermittent fasting should be initiated shortly after disease onset (8).

Fitzgerald et al. investigated safety, feasibility, weight loss and participant reported outcomes after 8 weeks of calorie restriction and intermittent fasting in individuals with MS. Both diets caused significant weight loss and increased emotional well-being (9).

KDs seem to be equally promising. KD was established as an alternative treatment for pharmaco-resistant childhood epilepsy in the 1920s (10). This was based on the observation that seizures stopped during fasting, but extended fasting periods in children were obviously not reasonable. Consequently, KD was developed to mimic biochemical effects caused by fasting (11). KDs also seem to improve symptoms of neurodegenerative diseases, such as Alzheimeŕs and Parkinsońs disease (12, 13). EAE mice on KD showed ameliorated disease progression, motor disability, hippocampal atrophy, lesion load, inflammation, and oxidative stress (14). In our own study we investigated feasibility, safety and health-related quality of life associated with a 6-month FD and KD intervention in individuals with relapsing-remitting MS (RRMS) (15). A single-arm, uncontrolled study reported improved metabolic and neurological outcomes after 6 months of a modified Atkins diet in individuals with RRMS (16, 17).

Based on this evidence, we designed a large-scale, long-term, randomized, controlled clinical study to investigate the effects of a FD and a KD compared to a standard healthy diet (SD) with anti-inflammatory focus in individuals with RRMS. We hypothesized that both FD and KD are superior to SD with respect to new cerebral lesions on cranial T2-weighted magnetic resonance imaging (MRI) after 18 months study interventions compared to baseline (18).

## Material and Methods

### Study design and published work

The Nutritional Approaches in MS (NAMS) study was a single center, randomized, controlled, parallel group study (ClinicalTrials.gov, NCT03508414). Participants were randomized to either SD, FD, or KD. The study was conducted at the Neuroscience Clinical Research Center of Charité – Universitätsmedizin Berlin from April 2017 to October 2021 (last visit). Beside the study protocol (18), an interim analysis on the effects of the dietary interventions on neuropsychiatric symptoms was published in the scope of a doctoral thesis (19).

### Participants

Main inclusion criteria were 1) definite diagnosis of RRMS according to the 2017 revised McDonald criteria (20) and 2) stable disease modifying therapy (DMT) or no DMT for at least six months prior to enrolment, 3) recent disease activity had to be confirmed, defined as at least one new lesion on cranial MRI or at least one clinical relapse within the last two years prior to enrolment, 4) Expanded Disability Status Scale score (EDSS) had to be < 4.5. Key exclusion criteria were start or change of DMT within six months before study start or during the study. The complete inclusion and exclusion criteria are listed in the published protocol (18). The study was approved by the institutional review board of Charité – Universitätsmedizin Berlin (EA1/200/16).

### Randomization

Participants were stratified according to DMT use (yes or no), sex (male or female), and T2 lesion load (low (≤ 15) vs. high (> 15)), to equally distribute potential confounder. Block randomization in treatment groups (1:1:1) was done by an external statistician, who was not involved in any study procedures. Whereas participants and study personnel could obviously not be blinded to the dietary intervention, outcome assessors (MRI, EDSS) were trained and blinded.

### Dietary interventions

Dietary interventions were instructed in 10 group sessions over 18 months. The group setting was chosen to encourage study adherence, explain dietary interventions, provide support, assess adverse events, and validly evaluate compliance. A detailed description of the interventions can be found in the study protocol (18). During the pandemic, groups sessions had to run virtually, temporarily.

The **SD group** followed a healthy, mainly vegetarian-focused diet according to the recommendations of the German Society for Nutrition (DGE). Participants were additionally instructed to adapt their omega-6 to omega-3 fatty acid intake to reach a ratio of 5:1 to set an anti-inflammatory focus. Total energy intake was not restricted. Following intermittent fasting patterns and/or any other diets was prohibited, thereby ensuring discrimination between interventions. The SD group, functionally interpreted as our control group, received an intervention for recruitment and ethical reasons (an 18-months follow-up of participants not receiving any intervention would have been unethical and hardly feasible).

The **FD group** fasted every 6 months for 7 days according to Buchinger (21). Participants in this group attended additional group sessions every second day during the 7-day fasts. Between these fasts, participants practiced 14:10 time-restricted eating on 6 days each week. Dietary intake recommendations were the same as in the SD group.

The **KD group** reduced their carbohydrate intake to 20-40 g/d and increased their fat intake to 70-80% of total energy intake. A focus on plant-based fats was advised. Participants were allowed to gradually increase their carbohydrate intake up to their individual limit (mostly at 40 g/d, maximum 50g/d) to maintain stable ketosis, defined as ≥ 0.5 mmol/L blood beta-hydroxybutyrate (BHB). For self-monitoring, participants received a standard hand-held ketone-meter (GlucoMen LX plus). Ketone values were discussed and followed-up during study visits and dietary group sessions by the study team. The state of ketosis and measuring of blood ketones allowed to evaluate compliance according to a biomarker.

### Outcome parameters and assessment methods

The primary endpoint and all secondary endpoints were measured at baseline, after 9 and 18 months. A detailed list of all study assessments with assessment time points can be found in the study protocol SPIRIT flow diagram (18).

### Primary outcome

Primary outcome was the number of new T2-hyperintense lesions found in cranial MRI after 18 months on FD or KD compared to SD.

### Secondary outcomes

#### MRI outcomes

**Brain atrophy** was determined by percentage of brain volume change (PBVC) at 18 months vs. baseline. To increase internal validity of the primary endpoint, the secondary MRI endpoint change of **lesion volume** was amended during the study. Enlarged and/or aggregated lesions may result in unchanged or even reduced lesion counts but signify disease progression. By controlling for lesion volume, data analysis was adjusted for this potential systematic error. All MRI scans were done in a 3-Tesla MRI scanner (Tim Trio, Siemens, Erlangen, Germany) and lesions were segmented by two experienced and blinded MRI technicians.

### Details on MRI Acquisition and Processing

Cerebral MRI scans were performed on two 3-Tesla (Siemens MAGNETOM Trio Tim and Prisma, Erlangen, Germany) scanner models, with 32-channel head coils. The Trio MRI (Scanner 1) protocol included: (1) a T1-weighted 3D magnetization prepared rapid gradient echo (MPRAGE) cerebral MRI (1mm isotropic resolution, repetition time (TR)=1900ms, time to echo (TE)=3.03ms, inversion time (TI)=900ms); (2) a 3D T2-weighted fluid-attenuated inversion recovery sequence (FLAIR) (1 mm isotropic resolution, TR=6000ms, TE=388ms, TI=1800ms). The Prisma MRI (Scanner 2) protocol included: (1) a 3D MPRAGE (1mm isotropic resolution, TR=1900ms, TE=3.03ms, TI=900ms), including the upper cervical cord; (2) a 3D FLAIR (1mm isotropic resolution, TR=6000ms, TE=390ms, TI=2100ms). For each participant at each timepoint, FLAIR images were co-registered with MPRAGE images, in which each follow-up MPRAGE was co-registered to the baseline MPRAGE in Montreal Neurological Institute (MNI) space using FMRIB’s Linear Image Registration Tool (FSL version 5.0.9). Whole brain T2-weighted hyperintense lesion masks were manually segmented from co-registered FLAIR scans using ITK-SNAP (www.itksnap.org) by 2 MRI technicians (over 10 years of MS research experience). Lesion counts and volumes extraction were performed on binary lesion masks using cluster and fslstats tools (https://fsl.fmrib.ox.ac.uk/fsl/fslwiki/Fslutils).

#### Participant-centered outcomes

**Fatigue** was assessed with the fatigue severity scale (FSS), with scores ≥ 4 indicating moderate-to-severe fatigue (22).

The presence of **depressive symptoms** was assessed with the Beck Depression Inventory-II (BDI-II, (23)), which is interpreted according to the following ranges: 0-8, no depressive symptoms; 9-13, minimal depressive symptoms; 14-19, mild depressive symptoms; 20-28, moderate depressive symptoms; 29-63, severe depressive symptoms (24).

**Cognition** (cognitive processing speed) was assessed with the oral Symbol Digit Modalities Test (SDMT, (25)).

**Hand grip strength** was assessed with a standard hand dynamometer (Jamar smart hand (26)) and **walking endurance** with the 6-min walk test (6MWT, (27)).

**Neurological–functional disability** was assessed with the EDSS and the Multiple Sclerosis Functional Composite (MSFC (28)), a scale for evaluating different parts and degrees of functional impairment in individuals with MS, i.e. gait speed (Timed 25-Foot Walk, T25FT), fine motor skills (9-Hole Peg Test, 9HPT) and cognitive function (Paced Auditory Serial Addition test, PASAT). The MSFC score was calculated using z-scores and compared to baseline values of the whole study population.

**Mental** and **physical health related quality of life** were assessed with the MSQol-54, an established tool to assess MS-related quality of life. It includes 54 items that can be summed up to 12 multi-item scales (physical health, physical role limitations, emotional role limitations, pain, emotional well-being, energy, social function, cognitive function, health perception, health distress, overall quality of life, sexual function) and two single items (change in health, sexual function satisfaction). Two summary scores – physical health composite and mental health composite – can be derived from a weighted combination of scale scores (29). MSQoL-54 scale scores were created using the Likert method by averaging items within the scales, and then row scores were linearly transformed into 0-100 scales. Higher values indicate better quality of life.

#### Anthropometry

Potential mediators of dietary effects were assessed via the following parameters: **body weight** was measured with a standard scale (seca GmbH and Co.KG) and **Body Mass Index (BMI)** was calculated (body weight/height (kg/m^2^)). **Body composition** was determined by bioelectrical impedance analysis (Biacorpus RX 4000, MEDI CAL healthcare GmbH, Karlsruhe, Germany).

#### Dietary intake

Macro- and micronutrient intake was calculated from food records, in which participants recorded the amounts of all consumed foods and drinks for four consecutive days (three week days and one weekend day), using the software OptiDiet PLUS 6.1 (GOE mbH, Linden, Germany) based on the German Nutrient Database.

#### Metabolism

Blood glucose, BHB, insulin, adiponectin, leptin, total cholesterol, LDL cholesterol, HDL cholesterol and triglycerides were measured after a 12-h overnight fast in the accredited laboratory (Labor Berlin, Charité Vivantes GmbH and Charité – Universitätsmedizin Berlin) according to standard procedures. Metabolic markers were assessed in order to investigate their potential of mediating effects of dietary interventions.

### Safety of dietary interventions

Safety was assessed by occurrence of (serious) adverse events, vital signs (blood pressure, heart rate), monitoring of body weight and markers for kidney (creatinine, uric acid, urea) and liver function (GPT, GOT, gamma-GT).

### Compliance with dietary interventions

Compliance for all participants was defined as attendance of at least 7 out of 10 group sessions over the study period. Food records at 0, 9 and 18 months allowed assessment of compliance with the different dietary recommendations. Compliance to the individual interventions was additionally defined as follows: SD, adherence to DGE recommendations; FD, attendance of dietary sessions during 7-day fasts; KD, 75% of study center assessed BHB concentrations ≥ 0.5 mmol/L.

### Data capture

Study data were collected and managed using REDCap electronic data capture tools hosted at Charité Berlin (30, 31).

### Samples size calculation and data analysis

Sample size calculation and data management procedures are published in the protocol publication (18). The primary endpoint was the number of new MRI lesions at 18 months assessed in the KD and FD group each in comparison to the SD group. Analysis of the primary endpoint was based on robust linear regression analysis in the ITT population, adjusted for baseline lesion load and lesion volume (to account for enlarged and/or aggregated lesions may result in unchanged or even reduced lesion counts but signify disease progression). In contrast to the initially planned ANCOVA analysis (18), the robust regression was preferred due to non-normally distributed data.

Sensitivity analysis of the primary outcome was done applying robust linear regression in the PP population. Secondary endpoint analysis was based on linear regressions and Wilcoxon signed rank tests in the FAS population.

We have decided not to impute for missing values in the outcome parameters. All available cases were considered. Analysis was conducted with the software R version 4.3.1 (R Core Team (2023). R: A language and environment for statistical computing. R Foundation for Statistical Computing, Vienna, Austria. URL https://www.R-project.org/) (32) and IBM SPSS statistics version 26.

## Results

During the recruitment period, 981 potential participants contacted the study center, of which 130 participants suited for screening and 105 were included (**Figure 1**).

**Figure 1.**
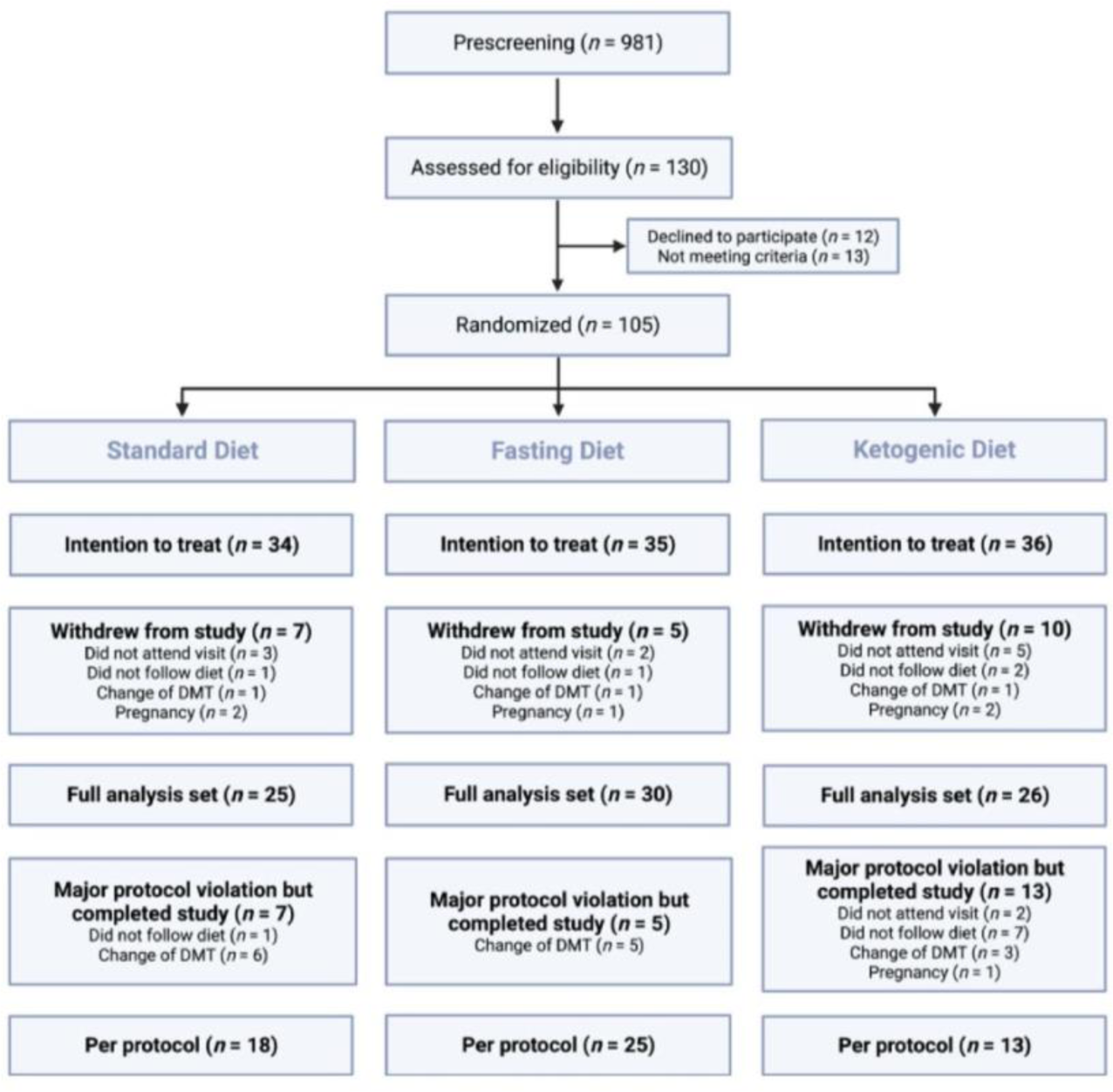
CONSORT diagram of tlie NAMS study. In the SD group: two MRIs were not done, which is w hy two more cases had to be excluded from the full analysis set. DMT. disease modifying therapy

Baseline characteristics are displayed in **Table 1**. The a priori sample size of 111 participants was not reached, as the dropout rate was higher than the initially estimated 10%. Dropout rates were 21% in the SD, 14% in the FD, 28% in the KD group (Figure 1).

**Table 1:** Baseline characteristics of the NAMS intention-to-treat population.

|  |  | Total | SD | FD | KD |
| --- | --- | --- | --- | --- | --- |
| Number of participants (n) |  | 105 | 34 | 35 | 36 |
| Age (years) | Mean (StD) | 42 (9) | 43 (10) | 41 (9) | 41 (10) |
| Ethnicity (Caucasian) | n (%) | 100 (95) | 34 (100) | 34 (97) | 32 (89) |
| Gender (female) | n (%) | 88 (84) | 29 (85) | 28 (80) | 31 (86) |
| Disease duration since manifestation (years) | Median (IQR) | 8.0 (13.8-2.4) | 5.0 (13.0-2.1) | 8.9 (13.6-3.8) | 8.6 (16.1-3.0) |
| Disease duration since diagnosis (years) | Median (IQR) | 3.6 (10.3-1.3) | 2.1 (5.4-1.2) | 5.3 (11.1-2.1) | 3.8 (10.6-1.3) |
| EDSS score | Median (IQR) | 2.0 (2.5-1.5) | 2.0 (2.5-1.5) | 2.0 (2.1-1.5) | 2.0 (3.1-1.5) |
| Number of T2-lesions | Median (IQR) | 35 (75-19) | 36 (76-21) | 39 (67-23) | 31 (76-17) |
| Participants without disease-modifying therapy | n (%) | 45 (43) | 14 (41) | 15 (43) | 16 (44) |
| Body Mass Index (kg/m<sup>2</sup>) | Median (IQR) | 24.3 (26.6-22.2) | 25.1 (26.8-22.8) | 23.6 (26.3-21.6) | 23.7 (26.3-22.3) |
| 25-OH-Vitamin D3 (nmol/L) | Mean (StD) | 101.0 (37.1) | 105.1 (37.4) | 93.9 (34.1) | 104.2 (39.7) |
| Fatigue severity scale score | Median (IQR) | 3.7 (5.3-2.2) | 4.3 (5.6-1.8) | 3.8 (5.2-2.8) | 3.4 (4.4-2.2) |
| Beck depression inventory II score | Median (IQR) | 7 (12-3) | 7 (12-2) | 9 (13-5) | 6 (11-3) |
| Symbol digit modalities test score | Median (IQR) | 56 (63-49) | 54 (62-50) | 57 (62-50) | 57 (65-47) |

EDSS, expanded disability status scale; FD, fasting diet; IQR, interquartile range; KD, ketogenic diet; SD, standard diet; StD, standard deviation

There were no relevant differences in age and sex between the groups. Disease duration of the SD group was shorter than of the KD and FD group. However, EDSS score, lesion load and percentage of participants on DMT was comparable between groups. The SD group showed lower cognition (SDMT) scores (age-adjusted cut-off ≤ 55 implies cognitive impairment) and higher fatigues (FSS) scores (cut-off ≥ 4 implies moderate fatigue), compared to KD and FD group.

### Adherence, safety, and tolerability of the diets

Eighty-one participants completed the study in the Full Analysis Set (FAS) population, in which all exploratory analyses of secondary endpoints were performed.

All diets were well tolerated, and adverse events were mostly mild and transient (total, n = 311; SD, n = 77; FD, n = 115; KD, n = 119). The portion of the groups and types of adverse events can be found in **Suppl. table 1**. Serious adverse events were not related to the diets (total, n = 11; SD, n = 2; FD, n = 2; KD, n = 7; **Suppl. table 2**).

Attendance rates of dietary sessions were 88%, 100% and 77% in the SD, FD, and KD group, respectively. In the KD group, plasma BHB was ≥ 0.5 mmol/L in 62% of participants at 9 months and in 35% at 18 months, with median concentrations ≥ 0.5 mmol/L, indicating nutritional ketosis on group level at both time points (**Figure 2A**).

**Figure 2.**
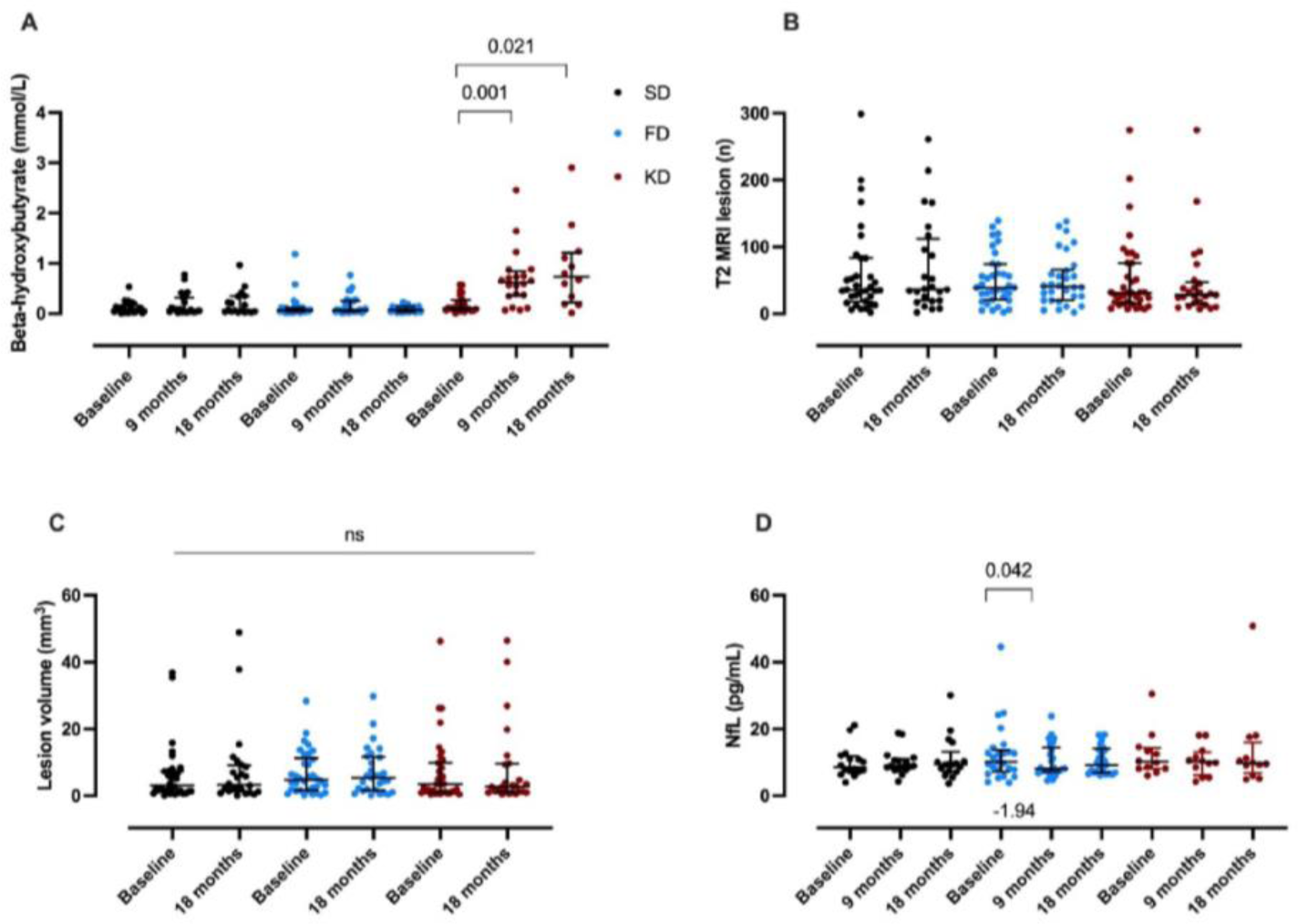
Adherence, imaging, and neurodegeneration markers in the standard diet (SD. black circles), fasting diet (FD. blue circles) and ketogenie diet (KD. red circles) of the NAMS study. A) Blood beta-hvdroxybutyrate concentrations confirmed nutritional ketosis in the KD group (FAS population) B) No change of the number of new MRI lesions (primary endpoint. ITT population) C) No change of MRI lesion volume (ITT population). **D)** Lower NIL concentrations at 9 months in the FD group (FAS population). A-C. data as median and interquartile range, p values by Wilcoxon signed-rank test. **D.** data as mean and standard deviation, p values by t-test

Food records revealed no relevant macro- or micronutrient intake deficiencies in the FD and KD group. The KD group reduced their carbohydrate intake from 40% at baseline to 13% and 20% of energy intake at 9 and 18 months, respectively. Their fat intake increased from 41% at baseline to 65% and 58% of energy intake at 9 and 18 months, respectively. Fiber intake increased in the SD group at 9 months vs. baseline by (+9 (18) g/d up to 33 (19) g/d, p = 0.028) and was higher than in the FD (20 (11) g/d) and the KD group (25 (8) g/d). At 18 months, fiber intake in the FD group (22 (10) g/d) was still lower than in the SD (30 (10) g/d) and KD group (27 (10) g/d). Comprehensive overviews on dietary intake (macro and micronutrient intake) in all participants and diet groups are displayed in **Suppl. table 6-9**.

#### Primary endpoint

Primary outcome analysis was applied in the ITT population. Treatment effects have been estimated from a robust linear regression with the number of new lesions as response, treatment group as fixed effect and adjusted for baseline number of T2-lesions and lesion volume. Treatment effects were estimated from Generalized Estimation Equations which show no difference between the groups regarding new number of T2 lesions at 18 months. Lesion volume influenced the number of new lesions at 18 months. (**Table 2).**

**Table 2.** Robust linear regression for number of new lesions at 18 months (ITT population)

| Variable | Unstandardized B | Std. Error | z value | p-value |
| --- | --- | --- | --- | --- |
| **Primary analysis (ITT)** |  |  |  |  |
| KD (SD as reference) | 0.305 | 1.205 | 0.253 | 0.801 |
| FD (SD as reference) | 0.809 | 1.171 | 0.691 | 0.492 |
| Lesion volume change (mm<sup>3</sup>) | -1.262 | 0.550 | -2.297 | 0.024 |
| Baseline lesion load (n) | -0.003 | 0.011 | -0.259 | 0.797 |

#### Secondary endpoints

All secondary endpoints were analyzed in the **FAS population** unless explicitly mentioned otherwise.

### Stable markers of disease progression

The number of new T2 lesions after 18 months vs. baseline did not change in any of the groups (primary endpoint; SD 0 (0-(−1)), FD 0 (2-0), KD 0 (2-0)) (Figure 2B). Sensitivity analysis investigated the per-protocol (PP) population and only emphasized significant predictive value for baseline lesion load (**Table 3**). In line with this, lesion volume remained stable over 18 months in all three groups (Figure 2C).

**Table 3.**
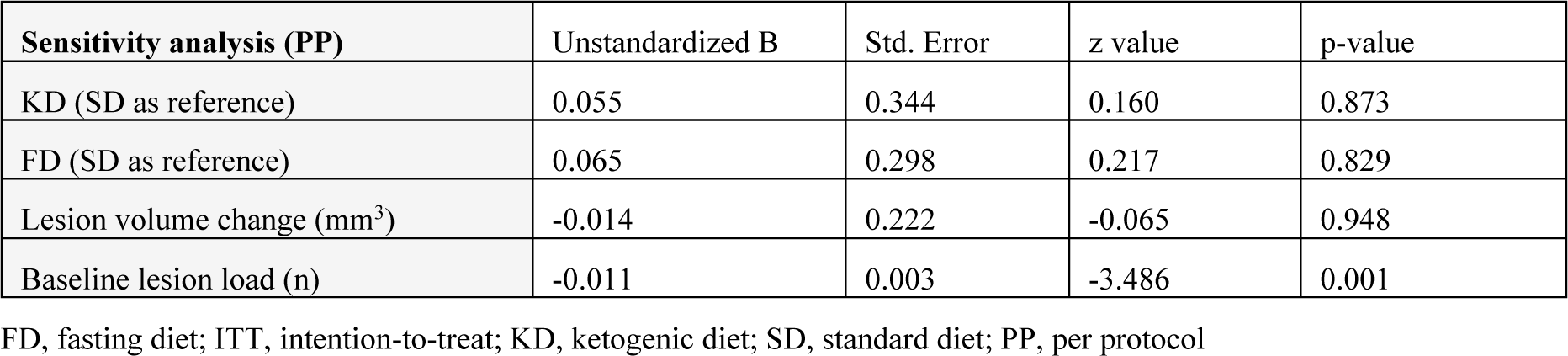
Robust linear regression for number of new lesions at 18 months (PP population)

| **Sensitivity analysis (PP)** | Unstandardized B | Std. Error | z value | p-value |
| --- | --- | --- | --- | --- |
| KD (SD as reference) | 0.055 | 0.344 | 0.160 | 0.873 |
| FD (SD as reference) | 0.065 | 0.298 | 0.217 | 0.829 |
| Lesion volume change (mm<sup>3</sup>) | -0.014 | 0.222 | -0.065 | 0.948 |
| Baseline lesion load (n) | -0.011 | 0.003 | -3.486 | 0.001 |

FD, fasting diet; ITT, intention-to-treat; KD, ketogenic diet; SD, standard diet; PP, per protocol

In addition, there was a significant predictive value for female sex and lesion volume change regarding the number of new lesions at 18 months in the FAS population (data not shown), but no influence of the dietary interventions. Percentage of brain volume change did not differ between the groups after 18 months study interventions (data not shown).

Serum neurofilament light chain (NfL) decreased by −1.94 ((−0.80)-0.01) pg/mL at 9 months in the FD group (Figure 2D) but not 18 months compared to baseline. NfL concentrations at 18 months correlated positively with EDSS score (Spearman correlation, r = 0.370; p = 0.006) and MRI lesion count (Spearman correlation, r = 0.394; p = 0.003) in a pooled analysis of all groups.

Participants reported 29 clinical relapses during the study (SD, 7 (28%); FD, 9 (30%); KD, 13 (50%)). Differences were not significant according to Chi-square tests (KD vs. FD vs. SD, p = 0.184; KD vs. SD, p = 0.186; FD vs. SD, p = 1; KD vs. FD, p = 0.210).

### Stable neurological-functional outcomes

Disability status remained stable over 18 months in all groups, assessed by EDSS, MSFC, 6MWT and hand grip strength. In line with this, there was no relevant change in mental health and physical health related quality of life (**Suppl. table 3-5**).

### Stable and improved neuropsychiatric outcomes

FSS scores remained stable over 18 months in all groups (Suppl. tables 3-5). SDMT scores remained stable in the SD group, slightly increased in the FD group, and increased in the KD group at 18 months (**Figure 3A**). BDI-II scores were slightly lower at 9 months (Unstandardized B = - 3.7, p = 0.065), and lower at 18 months (Unstandardized B = - 4.3, p = 0.015) in the FD vs. the SD group and improved over time in the FD group (Figure 3B).

**Figure 3:**
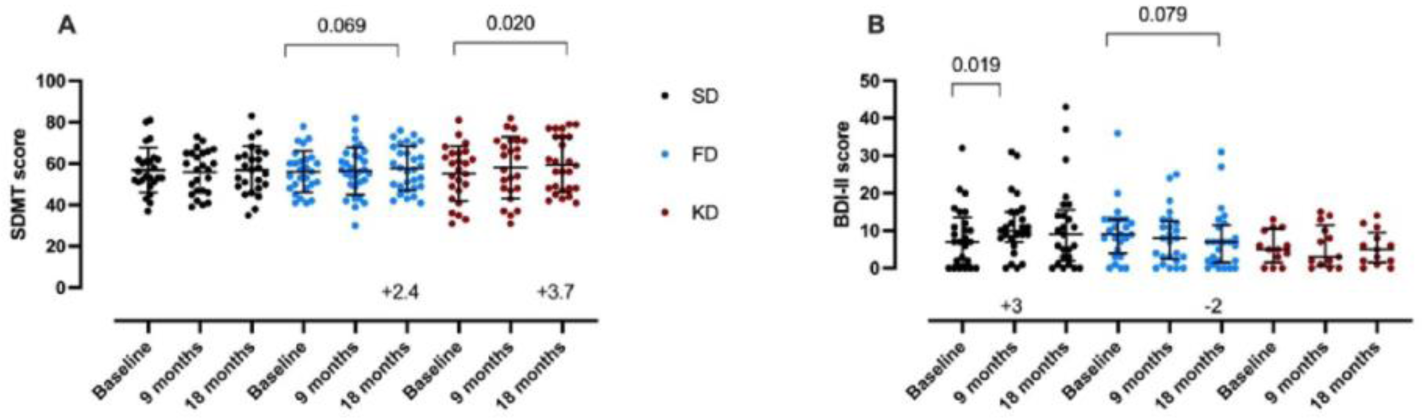
Ncuropsychiatric outcomes in the standard diet (SD. black circles), fasting diet (FD, blue circles) and kctogcnic diet (KD, red circles) of the NAMS study (FAS population) A) Improved cognition according to Symbol Digit Modalities Test (SDMT) at 18 months in the KD group. Data as mean and standard deviation, p value by t-test. B) Slightly improved depressive symptoms according to Beck Depression Inventory (BDI-II) in the FD group. Data as median and interquartile range, p values by Wilcoxon signed-rank test

### Improved cardiometabolic risk markers

Changes of cardiometabolic risk markers were most prominent at 9 months. Median **weight** losses at 9 months vs. baseline were 2.0 kg, 2.8 kg and 3.8 kg in the SD, FD, and KD group, respectively. Linear regression analysis showed that the KD (Unstandardized B = - 2.195, p = 0.039) and FD group (Unstandardized B = - 2.313, p = 0.022) had a lower body weight and BMI (KD, Unstandardized B = −0.9, p = 0.014) (FD, Unstandardized B = - 0.8, p = 0.015) compared to the SD group at 9 months, but not at 18 months. In all three groups, however, the **BMI** decreased significantly at 9 months (**Figure 4A**).

**Figure 4:**
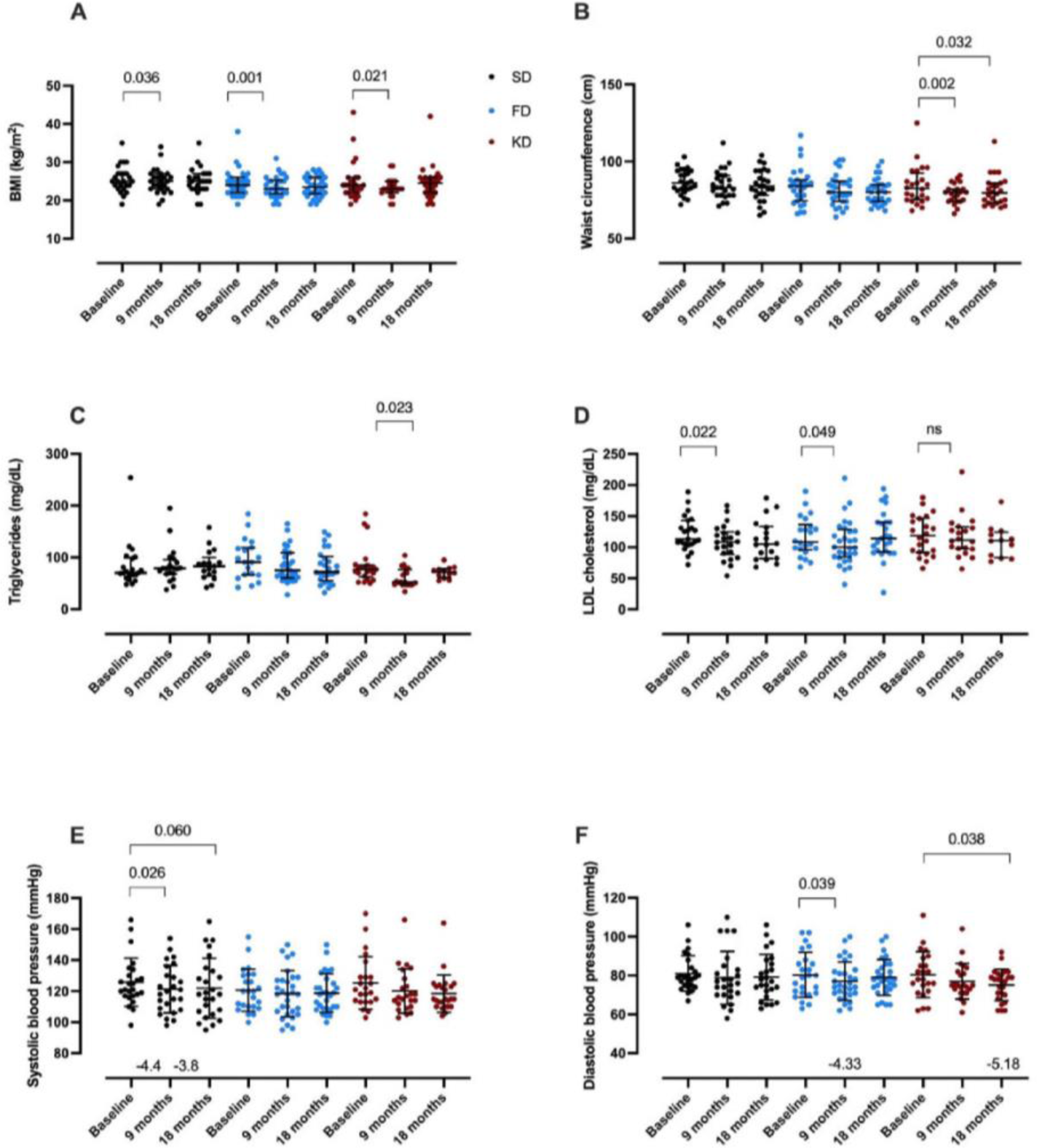
Improved cardiomctabolic risk markers in the standard diet (SD. blaek eireles). fasting diet (FD, blue eirelcs) and ketogenie diet (KD. red eireles) of the NAMS study (FAS population). A) Body mass index (BMI). **B)** waist circumference. **C)** triglycerides and D) LDL cholesterol Data as median and interquartile range, p values by Wileoxon signed-rank tests. **E)** Systolic and F) diastolic blood pressure Data as mean and standard deviation, p values by t-test

**Waist circumference** was 3 cm smaller at 18 months in the KD vs. the SD group (p = 0.044) and decreased within the KD group over time (Figure 4B). In line with this, **body fat mass** (Unstandardized B = - 1.653, p = 0.015) and **abdominal fat mass** (Unstandardized B = - 2.2, p = 0.008) were lower in the KD vs. the SD group at 9 months, but not at 18 months. In the KD group, abdominal fat mass decreased from 31% at baseline to 26% at 9 and 28% at 18 months.

**Body cell mass** (Unstandardized B = - 1.3, p = 0.022) and **fat free mass** (Unstandardized B = - 1.2, p = 0.047) were lower in the FD vs. the SD group at 9 months, but not 18 months.

**Leptin** concentrations were lower (Unstandardized B = - 3.0, p = 0.017) and **adiponectin** concentrations were higher (Unstandardized B = 2.2, p = 0.039) in the KD vs. the SD group at 9 months, but not at 18 months. Leptin decreased in the FD group (baseline: 9 (15-4) μg/L, 9 months: 6 (13-4) μg/L; Suppl. tables 3-5), however, not significant. In the KD group, Leptin decreased significantly by 3.5 (6.4) μg/L at 9 months (p = 0.044). Adiponectin significantly increased in the FD (+1.2 (2.3) μg/mL, p = 0.009) and with a trend in the KD group (+2.4 (5.4) μg/mL, p = 0.075).

In the KD group, **triglyceride** concentrations were 14 (21) mg/dL lower at 9 months vs. baseline (Figure 4C). In the SD group, **total cholesterol** decreased by 12 (19) mg/dL (p = 0.017; Suppl. tables 3-5)) and **LDL cholesterol** by 8 (14) mg/dL (p = 0.022) at 9 months vs. baseline (Figure 4D). In the FD group, LDL cholesterol decreased by a median of 8.5 (7.8-11.8) mg/dL (p = 0.049) comparing 9 months vs baseline (Figure 4D).

**Insulin** concentrations were lower at 9 months (Unstandardized B = - 1.7, p = 0.040) and 18 months (Unstandardized B = - 3.3, p = 0.001) in the FD group and at 18 months in the KD group (Unstandardized B = - 2.3, p = 0.058), each vs. the SD group (Suppl. tables 3-5).

Systolic blood pressure decreased in the SD group (**Error! Reference source not found.**), while diastolic blood pressure decreased at different time points in the KD and FD group (**Error! Reference source not found.**).

### Predictors and determinants for disease outcomes

Predictors for having at least one new lesion at 18 months could not be identified. BHB blood concentrations were not associated with improved cognition (linear regression analysis, p = 0.734). However, there was a trend for an inverse association between triglyceride concentrations at 9 months and the SDMT score at 18 months in the KD group only (linear regression analysis, adjusted R-square: 0.178, Unstandardized B: - 0.328, p = 0.064).

Additionally, there was an inverse correlation between diastolic blood pressure and SDMT scores at 18 months (Pearson correlation, r = - 0.396, p = 0.050) in the KD group, which could be reproduced as a trend in a linear regression (Unstandardized B = - 0.646, p = 0.054). Controlling for body weight, the trend remains (linear regression, Unstandardized B = - 0.626, p = 0.069).

The BDI-II score at 18 months was positively associated with body weight at 9 months in the FD group (trend, linear regression, Unstandardized B = 0.231, p = 0.054). Additionally, there was a positive correlation for leptin levels and BDI-II-scores at 18 months in the FD group (linear regression, Unstandardized B = 0.507, p = 0.043), a correlation that remained borderline significant when controlling for body weight (linear regression, Unstandardized B = 0.431, p = 0.059).

## Discussion

To our knowledge the NAMS study is the largest randomized-controlled study investigating in MS ketone-based diets (ketogenic and fasting diet) in comparison to a standard healthy diet. We show that these dietary approaches are safe with no relevant nutrient deficiencies, and only mild, transient adverse events. Repeated periodic fasting combined with time-restricted eating seemed to be more feasible than continuous reduction of carbohydrate intake, as evidenced by smaller dropout rates in the FD vs. KD group and less adherence in the KD at 18 months. Participants reported that the KD required drastic changes in daily life, much discipline, was time-consuming and socially less acceptable. Better adherence to time-restricted eating compared to continuous caloric restriction has been shown previously (33). Adverse events were more frequent in the FD and KD group, however, that may have been biased by increased awareness due to detailed explanation of possible side effects of KD and FD or more frequent meetings in the FD group.

We report no disease progression in terms of new cranial MRI lesions in any of the groups after 18 months study duration. Although a recent cohort study reported a relationship of pro-inflammatory diets with lesion volume and relapse rates (34), an anti-inflammatory dietary intervention did not show effects on MRI outcomes in a RCT (4). In contrast, a small randomized-controlled pilot study did show effects of 12-weeks intermittent caloric restriction on cortical volume and thickness (35). Even though we could not show an additional advantage of FD and KD, the complete lack of new lesions is a favorable result, especially considering the recruitment of people with MS with verified active disease (defined as one new lesion or relapse within the last two years prior to enrolment). In a comparable MS population, within 2 years, there was an increase of 17 new lesions in the placebo group compared to 3-4 new lesions under dimethyl fumarate (36). Noteworthy, 45% of our participants were on stable DMTs at study inclusion. This leads to a reduction in disease activity per se, which subsequently makes it more difficult to detect any influence on disease activity. However, including participants on different DMTs and no DMTs was unavoidable to ensure recruitment in a reasonable time frame and it certainly reflects real world conditions (18).

We found a decrease of NfL in the FD group at 9 months. As NfL concentrations correlated well with several MS disease outcomes (EDSS, lesions count, cognition) in our cohort and others (37, 38), this decrease may suggest decelerated axonal injury due to fasting (39). However, subjects with high BHB levels were reported to have a higher degree of improvement in serum NfL concentrations on KD in a study (40).

Elevated leptin concentrations are prevalent in MS (41), even independently of BMI (42), and are associated with reduced regulatory T-cells (43) as well as with more severe disability, also independently of BMI (44). In our KD group, leptin decreased, and adiponectin increased, in the KD and FD group. In addition, we found a positive association between leptin and depression scores in the FD group, indicating a positive effect of leptin reduction, which has been postulated previously (45).

Depressive symptoms, cognitive impairment and fatigue are highly prevalent in MS and may have detrimental effects on patient’s autonomy, socioeconomic status and quality of life (46). KDs have been shown to improve fatigue in MS, however, the lack of a control group (17) or very small sample sizes (47) limit inferences. Our study participants anecdotally described being less fatigued and more attentive while on KD. Despite, this was not reflected in the FSS scores of the overall groups. This discrepancy might be because the FSS rather queries physical fatigue than mental fatigue. Lower mental fatigue may be reflected by improved SDMT scores (+3.7 points at 18 months) in our KD group, a change considered clinically relevant (48).

Our diets had different, but overall beneficial effects on all measured blood lipids, which have been shown repeatedly to be associated with cognition in MS cohorts (49–52). In addition, NfL concentrations and HDL cholesterol were shown to be inversely correlated, suggesting metabolic alterations to be contributors to MS disease course (53).

There were slight effects on depressive symptoms in the FD group. A clinically relevant change of the BDI-II score is reported by a reduction of more than 17.5% (54). Our FD group improved by 22%, while our SD group deteriorated by 29%. This is in line with previous data showing improved domains of depressive symptoms due to fasting (55). In our FD group, body weight was positively associated with depression scores, indicating that this effect might have been mediated by weight reduction. Similar results were reported by Fitzgerald et al. (9).

Of note, the BDI-II does not seem to be best suitable for assessing depressive symptoms during dietary interventions, because it queries changes of appetite and sleep, which may be desirable side effects, e.g. in a KD (56, 57). Besides, when interpreting our results, it must be considered that depressive symptoms were only minimally prevalent at baseline in our cohort.

Our diets improved markers of cardiometabolic risk, such as body weight and composition, lipid profile, adipokines and blood pressure that are not only prevalent in MS, but also associated with an adverse disease course of MS (58–60). Especially hypertension affects grey matter atrophy and white matter damage (61). In line with Motl et al. (62), diastolic blood pressure, which was reduced by 5 mmHg at 18 months in our KD group, was inversely correlated with cognition scores. Vascular comorbidities have been shown to be associated with cognitive dysfunction in MS (63), suggesting a beneficial effect of blood pressure reduction due to dietary interventions.

We planned this study with 111 participants (expected dropout rate 10%) and included 105 participants with an actual dropout rate of 21%. We could not show a superiority of KD or FD diets compared to a healthy control diet: we suggest that the healthy standard diet of the control group has such a relevant influence on the disease activity, that it was not inferior to the ketone-based diets. However, for ethical reasons it was reasonable to offer the control group at least the standard dietary treatment, especially over a period of 18 months.

Furthermore, even though the overall study population is one of the largest nutritional intervention studies in MS, the group size of each intervention was quite small. Therefore, considering the heterogeneous disease course and use of DMTs of MS, our results need to be replicated in larger follow-up studies. However, in our study only a modest peripheral ketosis was measured and the full potential of ketone bodies as signaling molecules in the brain has probably not been achieved.

Of note, half of this study took place during the Covid-19 pandemic and was therefore severely affected by travel and contact restrictions, necessitating the change from in-person to online nutritional counselling. Additional effects of this turbulent time on our study outcomes cannot be ruled out. As dietary interventions are by nature unblinded, we aimed to minimize placebo effects by not communicating any interim results to participants.

The three group-setting based dietary approaches tested here may stabilize MS disease course and improve cardiometabolic health. We provide preliminary evidence that a ketogenic and a fasting diet may ameliorate cognition and depressive symptoms, respectively. Our study underlines the importance of dietary approaches as complementary treatment options.

## Data Availability

All data produced in the present study are available upon reasonable request to the authors

## Acknowledgements

We thank Claudia Messelhäußer and Bibiane Seeger-Schwinger for carrying out study visits and for laboratory pre-analytics, Susan Pikol and Cynthia Kraut for MRI T2-lesion segmentations, and Dania Schumann, Alexandra Prüß, Elektra Polychronidou for their engagement in carrying out the dietary sessions.

## Data sharing plan

Individual participant data that underlie the results of this article will be shared after deidentification on subject’s consent basis to researchers who provide a methodologically sound proposal on a related research question. Requests can be directed to.

## Authors’ contributions

JBS, MB, AM, FP, AMä designed the study. LSB, LF, DK, AP, DS, MB, CK, NS, AMä developed the interventional concept and accompanied the interventions during the course of the study. LSB, AM, FP and AMä procured funding. AK, JM and LSB applied statistical analyses. CC carried out MRI data management and extracted MRI metrics. LBS carried out study visits and data management. LSB and AMä wrote the manuscript. All authors read and approved the final version.

## Disclaimer

### Sources of support

Walter and Ilse Rose-foundation and Myelin project partially funded the study.

## Abbreviations

BDI: Beck depression inventory
BHB: Beta-hydroxybutyrate
BMI: Body Mass Index
DMT: Disease modifying therapy
EAE: Experimental autoimmune encephalomyelitis
EDSS: Expanded disability status scale
FAS: Full analysis set
FSS: Fatigue severity scale
FD: Fasting diet
ITT: Intention-to-treat
IQR: Interquartile range
KD: Ketogenic diet
MRI: Magnetic resonance imaging
MS: Multiple sclerosis
MSFC: Multiple sclerosis functional composite
NAMS: Nutritional approaches in multiple Sclerosis
NfL: Neurofilament light chain
PP: Per Protocol
QoL: Quality of life
RRMS: Relapsing-remitting multiple sclerosis
SD: Standard diet
SDMT: Symbol digit modalities test
StD: Standard deviation

## Supplement

**Supplemental table 1.**
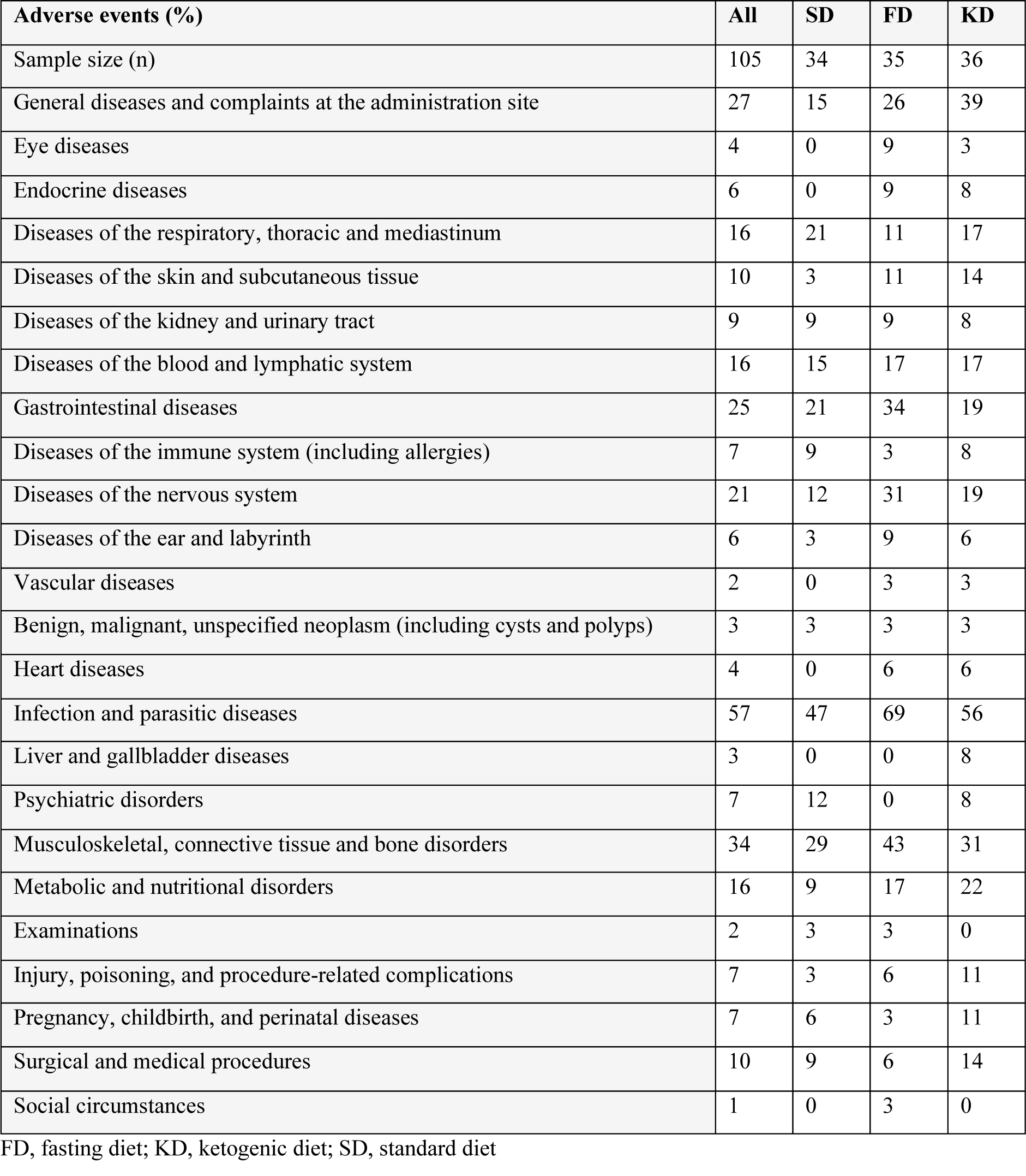
Adverse events in the Intention-to-Treat population of the NAMS study.

**Supplemental table 2.** Serious adverse events in the Intention-to-Treat population of the NAMS study.

| **Serious adverse events (n)** | **SD** | **FD** | **KD** |
| --- | --- | --- | --- |
| Imminently life-threatening | 1 | 0 | 1 |
| Hospital stay or extension | 1 | 1 | 5 |
| Coronavirus disease (COVID-19) | 0 | 1 | 1 |

FD, fasting diet; KD, ketogenic diet; SD, standard diet

**Supplemental table 3.** Outcome data of the NAMS study at baseline in the Full Analysis Set (FAS)

| FAS Baseline | All<br>(n) | mean<br>(StD) | median (Q3-Q1) | KD<br>(n) | mean<br>(StD) | median (Q3-Q1) | FD<br>(n) | mean<br>(StD) | median (Q3-Q1) | SD<br>(n) | mean<br>(StD) | median (Q3-Q1) |
| --- | --- | --- | --- | --- | --- | --- | --- | --- | --- | --- | --- | --- |
| Age (years) | 81 | 43<br>(10) | 44 (50-33) | 26 | 42<br>(10) | 43 (51-32) | 30 | 42<br>(9) | 43 (50-35) | 25 | 44<br>(10) | 46 (51-35) |
| EDSS score | 71 | 2.1<br>(1.0) | 2.0 (2.5-1.5) | 24 | 2.2<br>(1.2) | 2.0 (3.1-1.5) | 24 | 1.9<br>(0.9) | 2.0 (2.5-1.5) | 23 | 2.1<br>(0.8) | 2.0 (2.5-1.5) |
| T2 lesions, number (n) | 81 | 55.3<br>(58.1) | 34.0 (76.0-18.0) | 26 | 48.2<br>(58.3) | 29.0 (50.3-15.8) | 30 | 48.8<br>(38.1) | 39.0 (59.8-20.3) | 25 | 70.8<br>(74.7) | 36.0 (88.0-19.0) |
| T2 lesions, volume (mm<sup>3</sup>) | 81 | 6.7<br>(8.9) | 3.5 (8.4-1.2) | 26 | 7.0<br>(10.8) | 2.2 (7.7-1.0) | 30 | 6.4<br>(6.6) | 4.4 (9.6-1.5) | 25 | 6.9<br>(9.5) | 3.5 (7.3-1.5) |
| FSS score | 80 | 3.7<br>(1.8) | 3.7 (5.3-2.1) | 26 | 3.5<br>(1.6) | 3.5 (4.8-2.2) | 29 | 3.9<br>(1.8) | 3.9 (5.2-2.6) | 25 | 3.7<br>(1.9) | 3.8 (5.3-1.8) |
| BDI-II score | 81 | 9.0<br>(8.0) | 8.0 (12.0-3.0) | 26 | 8.7<br>(8.7) | 6.0 (10.8-3.0) | 30 | 9.5<br>(7.4) | 9.0 (13.0-5.3) | 25 | 8.5<br>(8.1) | 7.0 (12.0-2.0) |
| SDMT score | 77 | 56.0<br>(11.3) | 56.0 (62.0-49.0) | 25 | 55.1<br>(13.3) | 57.0 (65.0-47.0) | 27 | 56.0<br>(10.0) | 56.0 (61.5-49.0) | 25 | 56.8<br>(11.0) | 54.0 (62.0-51.0) |
| 6-MWT (min) | 79 | 558.3<br>(102.1) | 566.5 (616.2-502.5) | 25 | 556.0<br>(94.4) | 564.1 (608.1-487.6) | 29 | 559.2<br>(124.7) | 561.5 (638.1-502.7) | 25 | 559.6<br>(82.5) | 570 (609.8-510.8) |
| Hand grip strength (kg) | 78 | 33.6<br>(9.1) | 32.5 (37.1-27.6) | 26 | 35.2<br>(10.3) | 33.6 (38.6-27.6) | 28 | 32.0<br>(8.9) | 31.8 (36.4-25.9) | 24 | 33.9<br>(8.0) | 32.1 (36.6-27.8) |
| MSQOL54 Physical Health score | 80 | 69.6<br>(14.5) | 71.3 (78.5-63.4) | 25 | 68.3<br>(12.8) | 70.1 (77.6-65.9) | 30 | 70.7<br>(12.6) | 71.8 (76.6-65.8) | 25 | 69.5<br>(18.2) | 70.3 (85.2-54.2) |
| MSQOL54 Mental Health score | 80 | 68.1<br>(15.7) | 74.2 (79.9-57.7) | 25 | 65.7<br>(16.2) | 73.6 (79.8-57.5) | 30 | 68.5<br>(14.8) | 74.2 (78.8-59.2) | 25 | 67.0<br>(16.7) | 76.8 (83.1-57.8) |
| Average MSFC hpt dominant hand (sec) | 79 | 20.5<br>(3.4) | 19.8 (21.9-18.5) | 25 | 20.3<br>(3.9) | 19.3 (20.8-18.3) | 29 | 20.4<br>(2.6) | 19.9 (21.6-18.6) | 25 | 21.0<br>(3.7) | 20.8 (22.0-19.2) |
| Average MSFC t25fw (sec) | 78 | 4.3<br>(0.7) | 4.2 (4.7-3.8) | 25 | 4.3<br>(0.7) | 4.1 (4.8-3.9) | 28 | 4.3<br>(0.8) | 4.4 (4.7-3.9) | 25 | 4.1<br>(0.6) | 4.1 (4.5-3.7) |
| Z Score t25fw | 78 | 0.0<br>(1.1) | 0.1 (0.6-(-0.5)) | 25 | -0.1<br>(1.1) | 0.2 (0.4-(-0.9)) | 28 | -0.1<br>(1.2) | -0.1 (0.6-(-0.7)) | 25 | 0.2<br>(0.9) | 0.2 (0.8-(-0.3)) |
| **Z Score PASAT** | 78 | 0.1<br>(0.8) | 0.3 (0.8-(-0.4)) | 24 | 0.2<br>(0.7) | 0.2 (0.825-(-0.3)) | 29 | -0.2<br>(0.8) | -0.3 (0.6-(-0.6)) | 25 | 0.4<br>(0.7) | 0.6 (0.9-0.2) |
| **Z Score hpt** | 79 | -0.1<br>(1.0) | 0.0 (0.5-(-0.6)) | 25 | 0.1<br>(1.1) | 0.2 (0.56-(-0.2)) | 29 | -0.1<br>(0.8) | 0.0 (0.5-(-0.4)) | 25 | -0.2<br>(1.1) | -0.3 (0.3-(-0.7)) |
| **Overall Z Score** | 77 | 0.0<br>(0.7) | 0.2 (0.5-(-0.3)) | 24 | 0.1<br>(0.7) | 0.3 (0.6-(-0.2)) | 28 | -0.1<br>(0.7) | -0.1 (0.3-(-0.4)) | 25 | 0.1<br>(0.7) | 0.2 (0.6-(-0.1)) |
| **Disease duration from first manifestation (years)** | 81 | 9.2<br>(7.7) | 6.7 (14.8-2.3) | 26 | 10.4<br>(8.1) | 8.6 (16.5-2.9) | 30 | 9.0<br>(6.7) | 8.5 (14.6-4.2) | 25 | 8.2<br>(8.4) | 3.2 (12.7-1.9) |
| **Disease duration from first diagnosis (years)** | 80 | 6.1<br>(5.9) | 3.6 (10.1-1.4) | 26 | 6.4<br>(6.5) | 4.6 (9.6-1.3) | 30 | 7.0<br>(5.4) | 5.5 (12.0-2.1) | 24 | 4.5<br>(5.8) | 1.8 (5.1-1.1) |
| **BMI (kg/m<sup>2</sup>)** | 81 | 24.9<br>(4.1) | 24.3 (26.3-22.3) | 26 | 25.0<br>(5.2) | 23.7 (25.8-22.2) | 30 | 24.3<br>(3.7) | 23.7 (25.9-21.5) | 25 | 25.5<br>(3.4) | 25.0 (27.1-23.7) |
| **Body fat (%)** | 78 | 29.6<br>(8.1) | 29.7 (34.4-24.7) | 25 | 29.5<br>(9.9) | 29.1 (32.5-24.1) | 29 | 29.0<br>(6.8) | 29.3 (33.5-24.9) | 24 | 30.2<br>(7.7) | 32.3 (34.6-25.9) |
| **Total cholesterol (mg/dL)** | 67 | 192<br>(35) | 188 (212-173) | 22 | 187.6<br>(36.3) | 182 (205-156) | 22 | 195<br>(36) | 196 (214-176) | 23 | 193<br>(33) | 188 (206-179) |
| **LDL cholesterol (mg/dL)** | 67 | 119<br>(30) | 113 (142-97) | 22 | 119<br>(32) | 119 (145-93) | 22 | 117<br>(32) | 109 (132-97) | 23 | 122<br>(28) | 113 (137-106) |
| **HDL cholesterol (mg/dL)** | 67 | 70<br>(17) | 71 (80-57) | 22 | 69<br>(16) | 71 (79-54) | 22 | 74<br>(19) | 79 (83-57) | 23 | 69<br>(14) | 68 (79-62) |
| **Triglycerides (mg/dL)** | 62 | 88<br>(40) | 77 (100-66) | 22 | 85<br>(37) | 77 (84-66) | 19 | 96<br>(39) | 91 (118-67) | 21 | 85<br>(44) | 70 (98-66) |
| **Insulin (mU/L)** | 81 | 8.5<br>(5.1) | 7.0 (9.5-5.8) | 26 | 8.9<br>(6.1) | 6.5 (8.6-6.1) | 30 | 8.0<br>(4.5) | 7.0 (9.3-5.1) | 25 | 8.8<br>(4.7) | 7.4 (9.6-5.9) |
| **Leptin (µg/L)** | 75 | 11.3<br>(8.8) | 9.6 (15.0-5.1) | 22 | 11.8<br>(10.9) | 9.5 (14.3-5.2) | 28 | 10.3<br>(8.7) | 8.6 (14.2-4.2) | 25 | 11.9<br>(6.9) | 9.7 (17.3-6.9) |
| **Adiponectin (µg/mL)** | 79 | 9.5<br>(5.1) | 8.4 (12.1-5.6) | 24 | 10.1<br>(4.8) | 8.9 (13.2-6.3) | 30 | 9.1<br>(5.5) | 8.7 (11.4-4.5) | 25 | 9.4<br>(5.1) | 8.0 (11.7-6.3) |
| **Vitamin D (nmol/L)** | 79 | 101.6<br>(35.9) | 97.9 (119.6-79.5) | 25 | 103.2<br>(33.4) | 97.9 (117.3-86.6) | 29 | 93.8<br>(35.7) | 92.2 (119.5-70.1) | 25 | 108.9<br>(38.2) | 103.9 (125.7-82.1) |
| **Beta-hydroxybutyrate (mmol/L)** | 79 | 0.14<br>(0.18) | 0.09 (0.17-0.04) | 24 | 0.19<br>(0.17) | 0.11 (0.23-0.08) | 30 | 0.13<br>(0.23) | 0.07 (0.12-0.04) | 25 | 0.11<br>(0.12) | 0.07 (0.15-0.04) |
| **NfL (pg/mL)** | 55 | 11.5<br>(7.1) | 9.8 (12.8-7.3) | 12 | 12.3<br>(6.7) | 10.2 (14.3-8.1) | 25 | 12.5<br>(8.7) | 10.1 (13.7-7.2) | 17 | 10.0<br>(4.5) | 9.1 (11.6-7.3) |

FD, fasting diet; KD, ketogenic diet; SD, standard diet

**Supplemental table 4. O.**
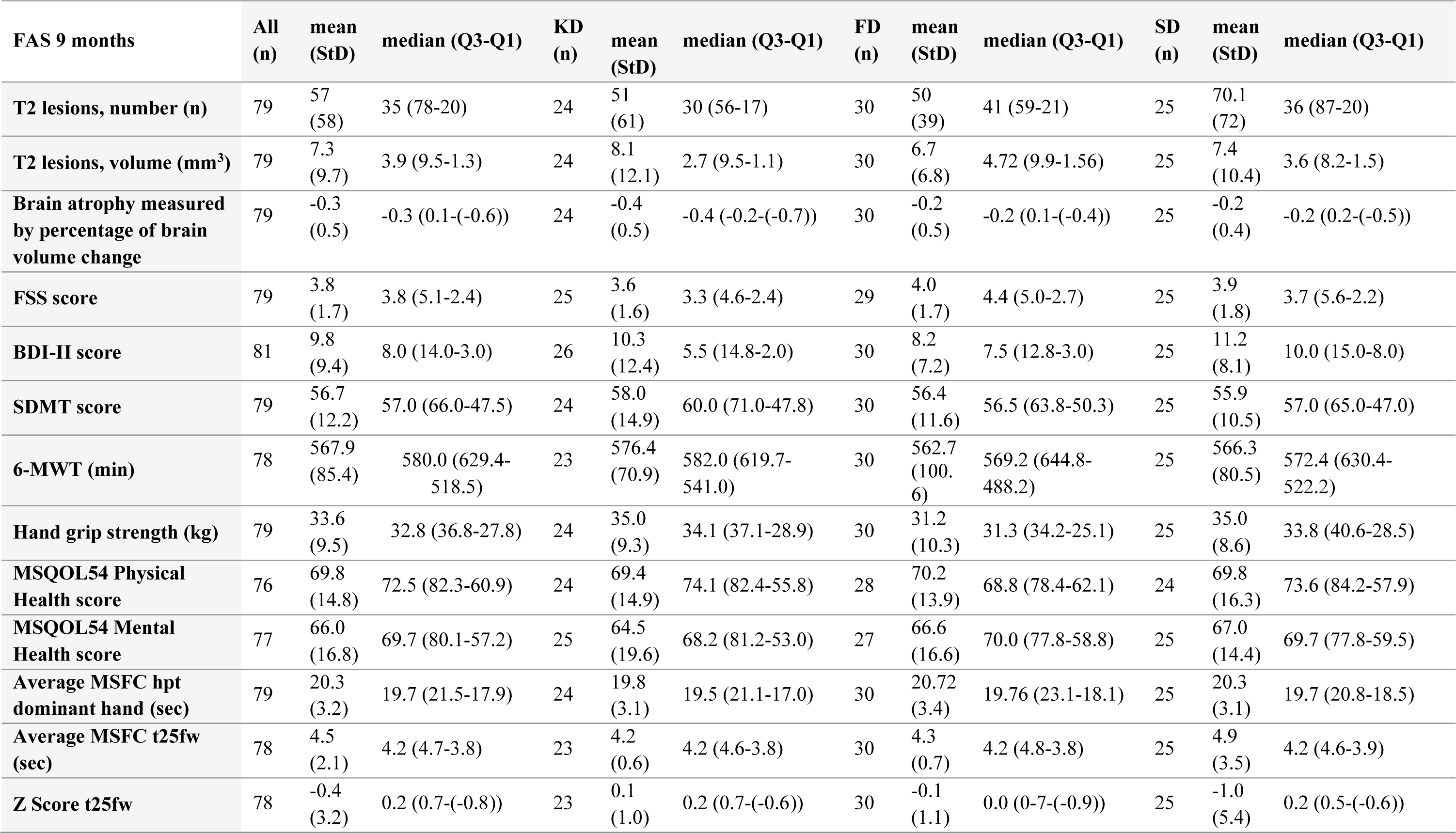
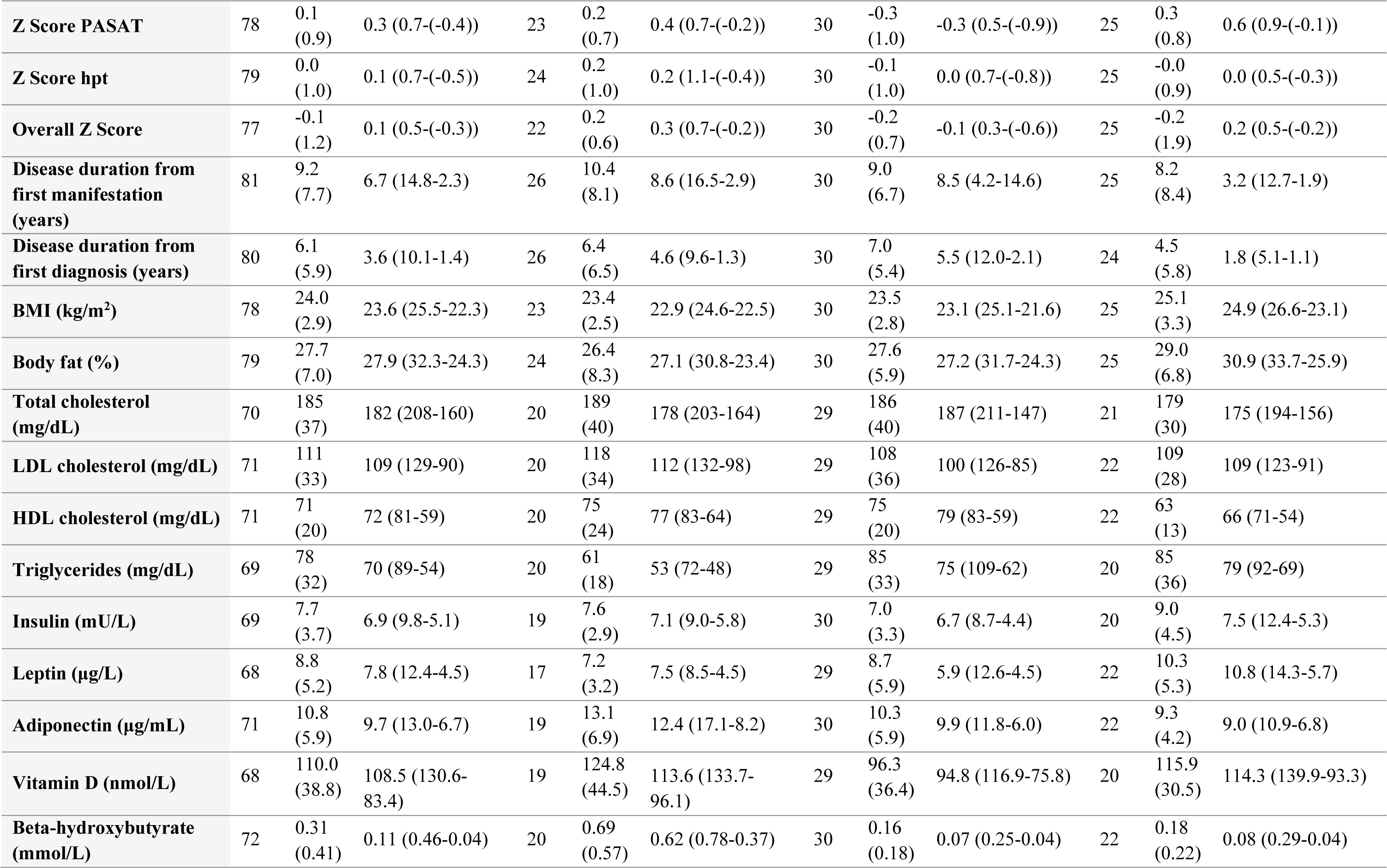
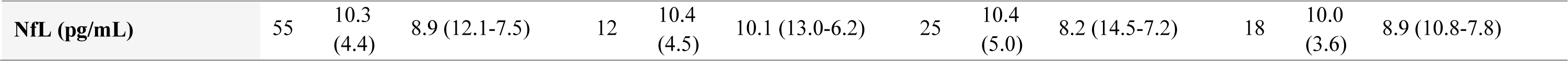
utcome data of the NAMS at 9 months in the Full Analysis Set (FAS)

**Supplemental table 5.** Outcome data of the NAMS at 18 months in the Full Analysis Set (FAS)

| **FAS 18 months** | **All**<br>**(n)** | **mean**<br>**(StD)** | **median (Q3-Q1)** | **KD**<br>**(n)** | **mean**<br>**(StD)** | **median (Q3-Q1)** | **FD**<br>**(n)** | **mean**<br>**(StD)** | **median (Q3-Q1)** | **SD**<br>**(n)** | **mean**<br>**(StD)** | **median (Q3-Q1)** |
| --- | --- | --- | --- | --- | --- | --- | --- | --- | --- | --- | --- | --- |
| **T2 lesions, number (n)** | 79 | 55<br>(57) | 35 (68-20) | 25 | 48<br>(59) | 28 (45-16) | 30 | 50<br>(39) | 41 (63-22) | 24 | 70<br>(71) | 37 (101-20) |
| **T2 lesions, volume (mm<sup>3</sup>)** | 79 | 7.5<br>(10.4) | 3.6 (9.5-1.3) | 25 | 8.0<br>(12.4) | 2.7 (9.4-1.2) | 30 | 7.0<br>(7.1) | 5.0 (11.1-1.6) | 24 | 7.8<br>(11.8) | 3.3 (8.8-1.4) |
| **Brain atrophy measured**<br>**by percentage of brain**<br>**volume change** | 79 | -0.6<br>(0.8) | -0.5 (-0.1-(-0.8)) | 25 | -0.7<br>(0.8) | -0.7 (-0.2-(-1.0)) | 30 | -0.4<br>(0.8) | -0.2 (0.2-(-0.7)) | 24 | -0.6<br>(0.6) | -0.5 (-0.2-(-0.7)) |
| **FSS score** | 81 | 3.9<br>(1.8) | 4.1 (5.3-2.2) | 26 | 3.8<br>(1.7) | 3.6 (5.1-2.7) | 30 | 4.1<br>(1.8) | 4.8 (5.6-2.7) | 25 | 3.6<br>(1.9) | 3.6 (4.9-1.8) |
| **BDI-II score** | 81 | 9.3<br>(9.7) | 7.0 (13.0-2.0) | 26 | 9.7<br>(9.8) | 6.0 (11.8-3.3) | 30 | 7.5<br>(7.7) | 7.0 (10.5-2.0) | 25 | 11.0<br>(11.4) | 9.0 (14.0-3.0) |
| **SDMT score** | 81 | 58.0<br>(11.8) | 57.0 (68.0-48.0) | 26 | 59.4<br>(13.3) | 57.5 (73.0-48.0) | 30 | 57.7<br>(10.7) | 57.5 (67.3-49.0) | 25 | 56.8<br>(11.6) | 57.0 (64.0-49.0) |
| **6-MWT (min)** | 81 | 559.2<br>(90.4) | 578.0 (625.3-485.0) | 26 | 552.0<br>(96.5) | 576.3 (600.6-509.4) | 30 | 558.7<br>(87.2) | 561.3 (620.9-502.3) | 25 | 567.4<br>(91.2) | 593.5 (636.0-517.3) |
| **Hand grip strength (kg)** | 78 | 33.8<br>(9.4) | 32.6 (37.1-27.8) | 24 | 35.7<br>(38.5-28.2) | 36.1 (38.5-28.2) | 29 | 31.5<br>(10.1) | 31.2 (34.6-25.9) | 25 | 34.7<br>(7.8) | 33.6 (37.6-28.5) |
| **MSQOL54 Physical**<br>**Health score** | 79 | 70.4<br>(16.8) | 76.3 (81.9-59.7) | 25 | 69.4<br>(15.5) | 75.3 (80.5-56.6) | 29 | 72.2<br>(14.6) | 76.0 (81.9-66.5) | 25 | 69.3<br>(20.6) | 79.3 (84.2-50.2) |
| **MSQOL54 Mental**<br>**Health score** | 81 | 67.3<br>(16.6) | 71.3 (80.5-58.9) | 26 | 65.9<br>(17.1) | 70.2 (80.7-52.5) | 30 | 67.8<br>(15.2) | 70.0 (80.1-58.9) | 25 | 68.1<br>(18.2) | 73.7 (80.5-62.2) |
| **Average MSFC hpt**<br>**dominant hand (sec)** | 80 | 20.1<br>(3.3) | 19.5 (21.1-17.7) | 25 | 19.4<br>(3.1) | 18.9 (20.2-17.7) | 30 | 20.9<br>(3.5) | 20.7 (21.8-18.5) | 25 | 19.8<br>(3.2) | 19.3 (21.0-17.7) |
| **Average MSFC t25fw (sec)** | 81 | 4.4<br>(0.7) | 4.2 (4.7-3.9) | 26 | 4.4<br>(0.9) | 4.2 (4.8-3.9) | 30 | 4.4<br>(0.6) | 4.3 (4.8-3.9) | 25 | 4.2<br>(0.5) | 4.1 (4.5-3.9) |
| **Z Score t25fw** | 81 | -0.2<br>(1.1) | 0.1 (0.6-(-0.7)) | 26 | -0.2<br>(1.4) | 0.2 (0.6-(-0.8)) | 30 | -0.2<br>(1.0) | -0.1 (0.4-(-0.8)) | 25 | -0.0<br>(0.8) | 0.1 (0.6-(-0.4)) |
| **Z Score PASAT** | 80 | 0.2<br>(1.0) | 0.4 (0.9-(-0.1)) | 25 | 0.2<br>(0.9) | 0.4 (0.8-0.1) | 30 | -0.1<br>(1.1) | 0.2 (0.7-(-0.6)) | 25 | 0.4<br>(0.8) | 0.7 (1.0-0.2) |
| **New Z Score hpt** | 80 | 0.1<br>(1.0) | 0.1 (0.8-(-0.4)) | 25 | 0.3<br>(1.0) | 0.3 (0.9-(-0.1)) | 30 | -0.2<br>(1.0) | -0.3 (0.6-(-0.6)) | 25 | 0.2<br>(1.0) | 0.2 (0.8-(-0.4)) |
| **Overall Z Score** | 79 | 0.1<br>(0.7) | 0.3 (0.5-(-0.3)) | 24 | 0.2<br>(0.8) | 0.4 (0.6-0.1) | 30 | -0.2<br>(0.7) | 0.0 (0.3-(-0.5)) | 25 | 0.2<br>(0.6) | 0.3 (0.6-(-0.2)) |
| **Disease duration from first manifestation (years)** | 81 | 9.2<br>(7.7) | 6.7 (14.8-2.3) | 26 | 10.4<br>(8.1) | 8.6 (16.5-2.9) | 30 | 9.0<br>(6.7) | 8.5 (14.6-4.2) | 25 | 8.2<br>(8.4) | 3.2 (12.7-1.9) |
| **Disease duration from first diagnosis (years)** | 80 | 6.1<br>(5.9) | 3.6 (10.1-1.4) | 26 | 6.4<br>(6.5) | 4.6 (9.6-1.3) | 30 | 7.0<br>(5.4) | 5.5 (12.0-2.1) | 24 | 4.5<br>(5.8) | 1.8 (5.1-1.1) |
| **BMI (kg/m<sup>2</sup>)** | 81 | 24.4<br>(3.5) | 24.3 (26.0-22.2) | 26 | 24.5<br>(4.4) | 24.5 (25.7-22.3) | 30 | 23.5<br>(2.6) | 23.4 (25.5-21.7) | 25 | 25.2<br>(3.3) | 24.8 (26.9-23.1) |
| **Body fat (%)** | 77 | 28.4<br>(7.4) | 30.6 (32.8-23.6) | 23 | 27.8<br>(9.8) | 30.8 (33.4-21.1) | 29 | 27.9<br>(5.7) | 28.5 (31.5-23.6) | 25 | 29.6<br>(6.6) | 31.2 (34.5-26.5) |
| **Total cholesterol (mg/dL)** | 55 | 190<br>(40) | 187 (211-162) | 12 | 186<br>(39) | 180 (211-156) | 25 | 197<br>(42) | 190 (228-170) | 18 | 182<br>(39) | 182.5 (200-152) |
| **LDL cholesterol (mg/dL)** | 55 | 115<br>(34) | 110 (135-91) | 12 | 109<br>(27) | 111 (122-86) | 25 | 120<br>(38) | 114 (140-94) | 18 | 111<br>(33) | 105 (130-84) |
| **HDL cholesterol (mg/dL)** | 55 | 71<br>(19) | 69 (83-57) | 12 | 74<br>(25) | 66 (92-54) | 25 | 75<br>(17) | 71 (88-64) | 18 | 62<br>(16) | 63 (69-53) |
| **Triglycerides (mg/dL)** | 54 | 80<br>(28) | 75 (90-63) | 12 | 71<br>(11) | 72 (78-65) | 25 | 82<br>(32) | 72 (98-58) | 17 | 85<br>(30) | 83 (90-67) |
| **Insulin (mU/L)** | 53 | 7.7<br>(3.8) | 6.8 (9.3-5.3) | 11 | 7.4<br>(2.4) | 7.1 (7.4-6.2) | 24 | 6.3<br>(2.7) | 5.7 (7.0-4.6) | 18 | 9.8<br>(4.7) | 9.3 (12.7-5.7) |
| **Leptin (µg/L)** | 52 | 9.7<br>(6.5) | 9.0 (12.4-5.0) | 10 | 9.0<br>(5.4) | 9.3 (12.3-4.1) | 25 | 10.0<br>(6.5) | 9.6 (13.7-5.0) | 17 | 9.6<br>(7.5) | 8.7 (11.1-4.9) |
| **Adiponectin (µg/mL)** | 53 | 9.8<br>(4.3) | 8.9 (12.2-7.0) | 11 | 11.0<br>(4.7) | 9.5 (14.9-7.6) | 25 | 9.7<br>(4.1) | 9.5 (11.2-7.0) | 17 | 9.4<br>(4.5) | 7.6 (12.5-6.4) |
| **Vitamin D (nmol/L)** | 53 | 114.7<br>(42.7) | 107.6 (143.5-85.0) | 11 | 126.4<br>(40.0) | 129.1 (152.8-88.3) | 24 | 107.5<br>(47.4) | 93.8 (123.5-77.7) | 18 | 117.2<br>(37.9) | 119.4 (139.1-92.6) |
| **Beta-hydroxybutyrate (mmol/L)** | 54 | 0.31<br>(0.51) | 0.12 (0.31-0.04) | 12 | 0.90<br>(0.81) | 0.74 (1.14-0.30) | 24 | 0.09<br>(0.06) | 0.07 (0.12-0.03) | 18 | 0.2<br>(0.25) | 0.07 (0.31-0.04) |
| **NfL (pg/mL)** | 55 | 11.3<br>(7.2) | 9.4 (13.8-7.0) | 12 | 13.5<br>(12.5) | 9.8 (16.0-6.8) | 25 | 10.6<br>(4.0) | 9.2 (14.1-6.9) | 18 | 10.9<br>(6.2) | 9.4 (11.7-7.3) |

**Supplemental table 6:**
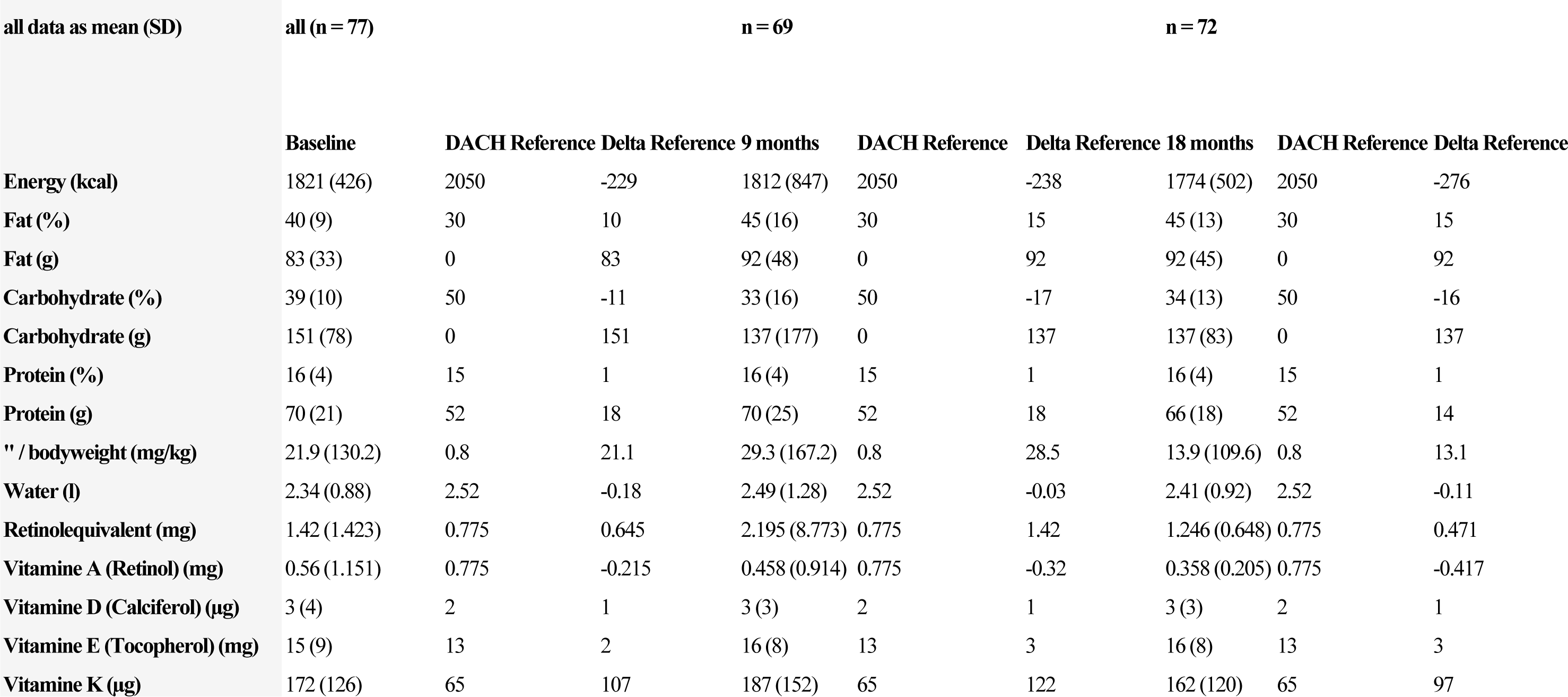
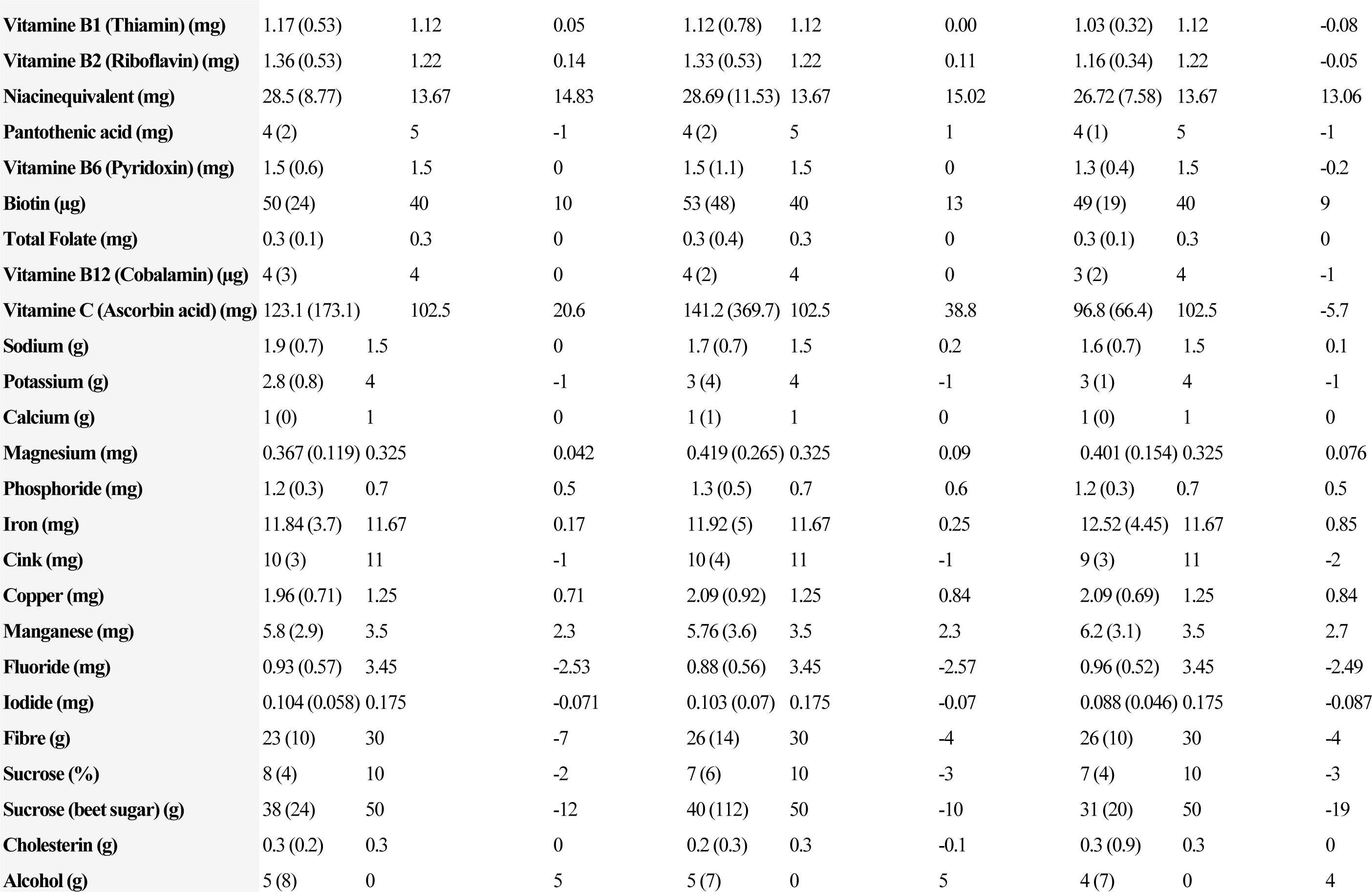
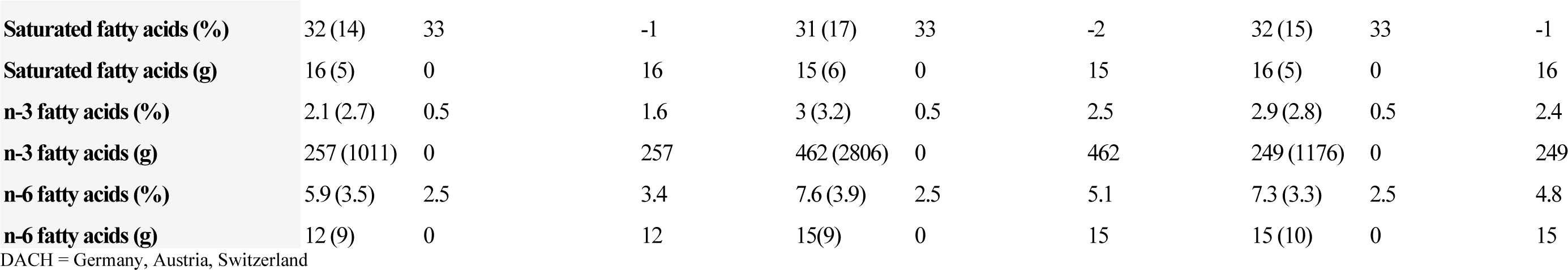
Dietary intake data for all participants.

**Supplemental table 7:**
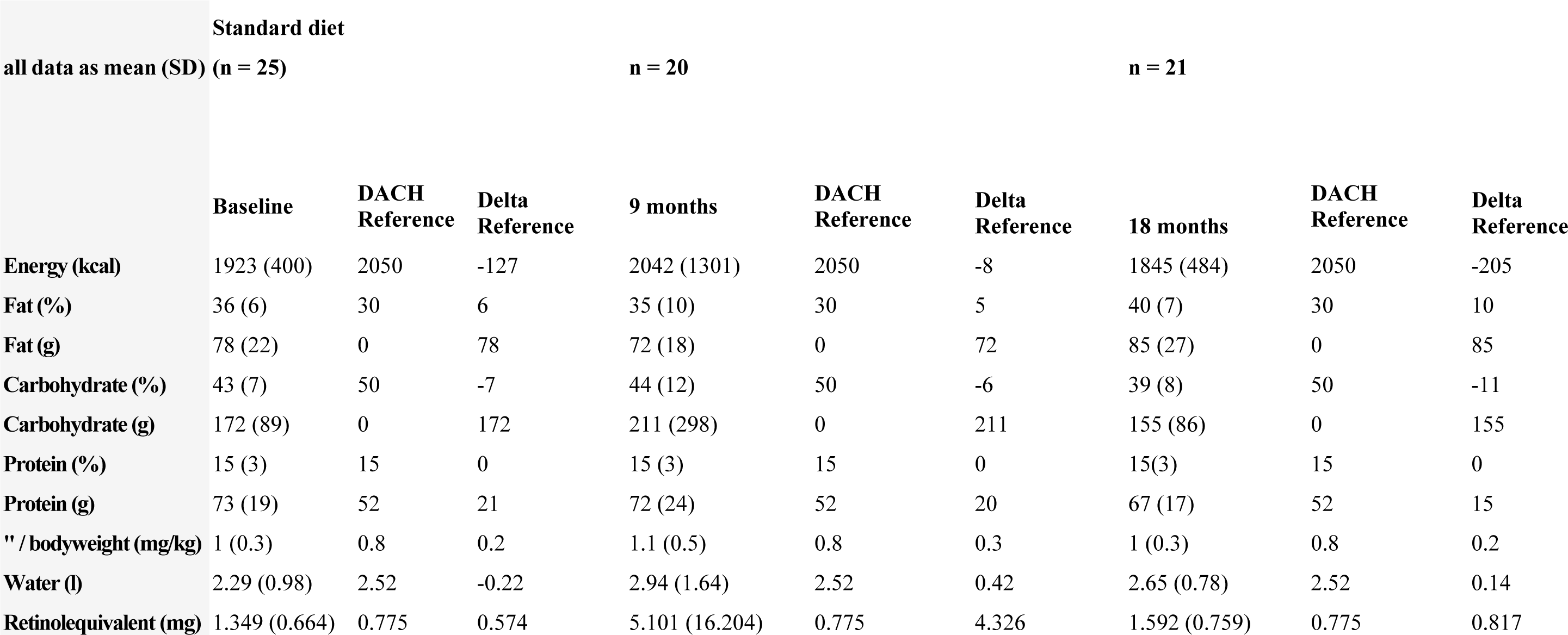
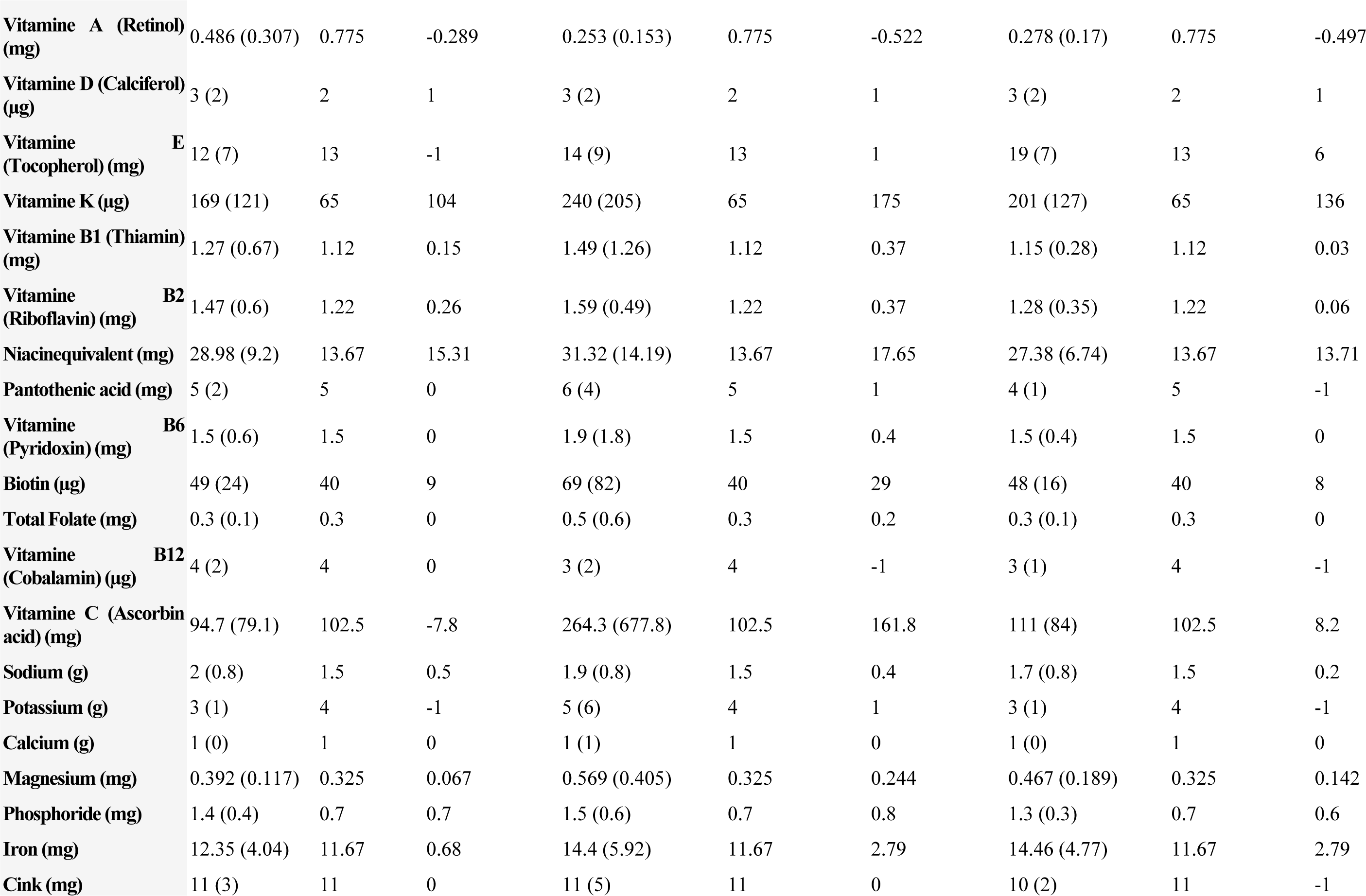
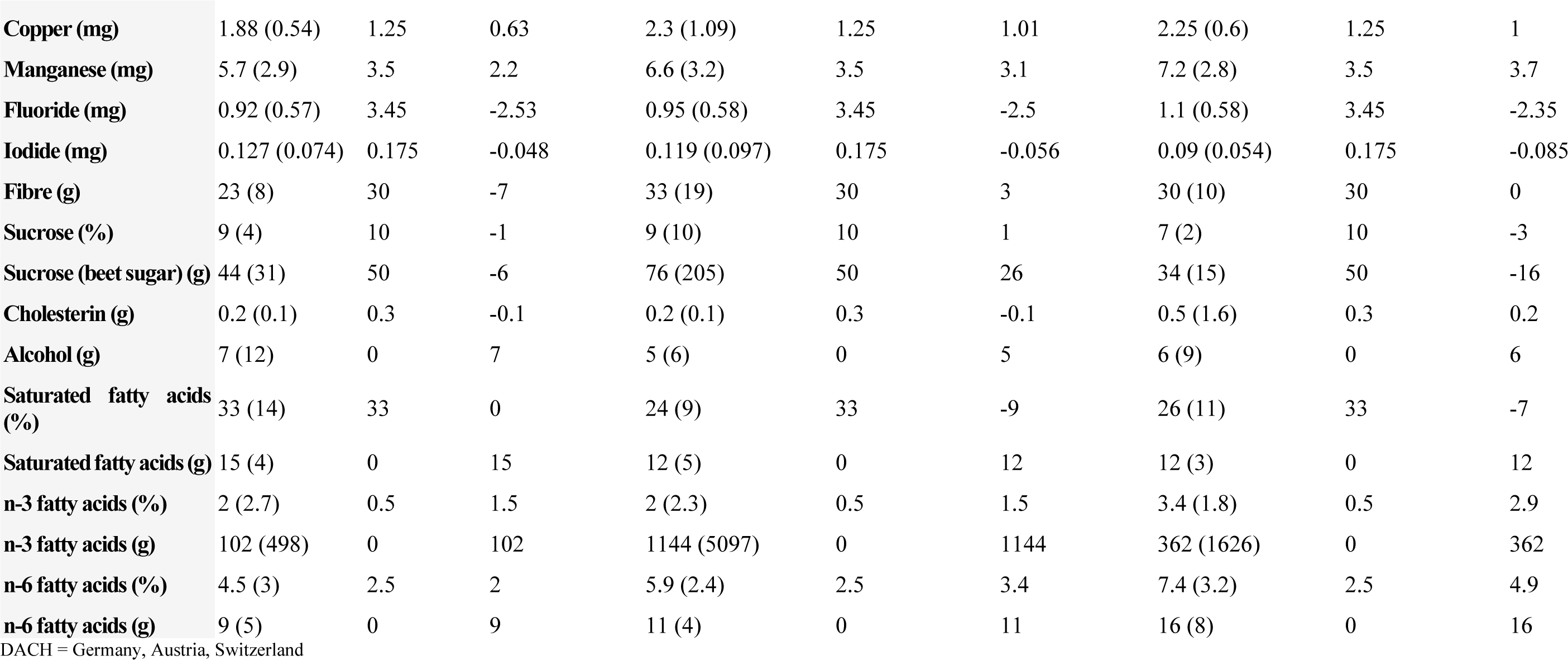
Dietary intake data for standard diet group.

**Supplemental table 8:**
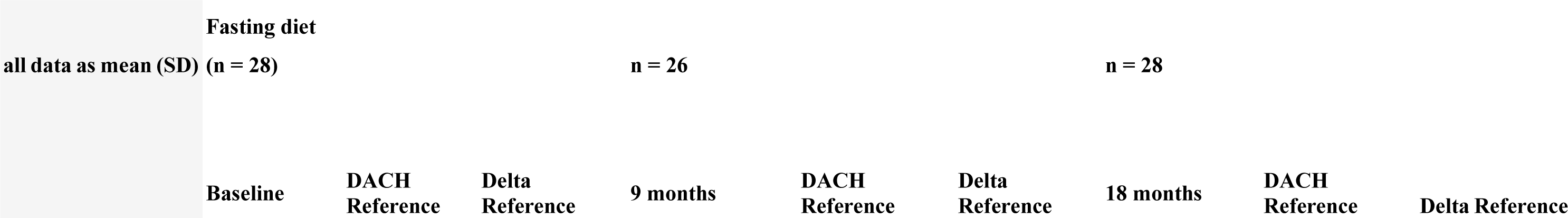
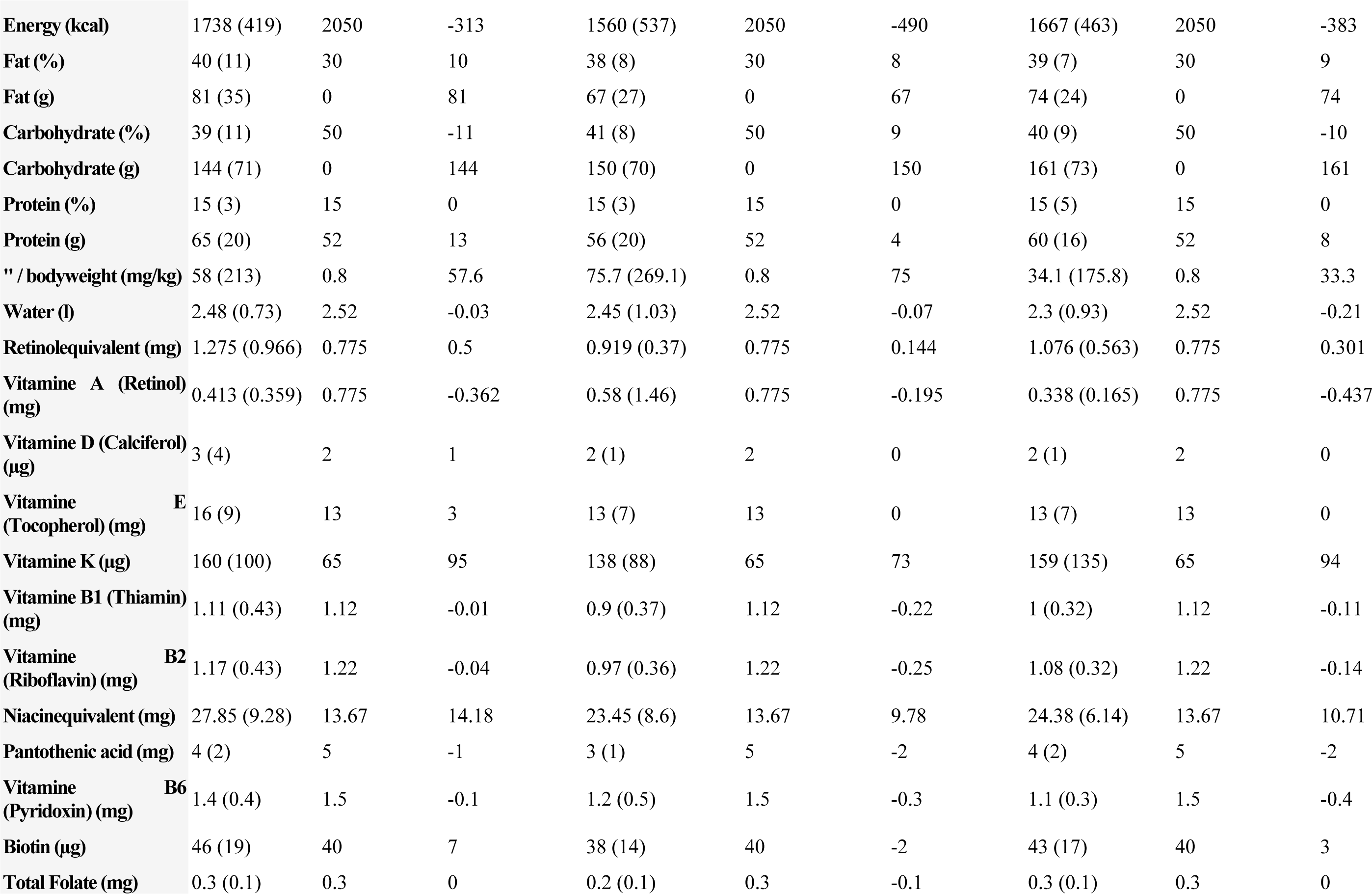
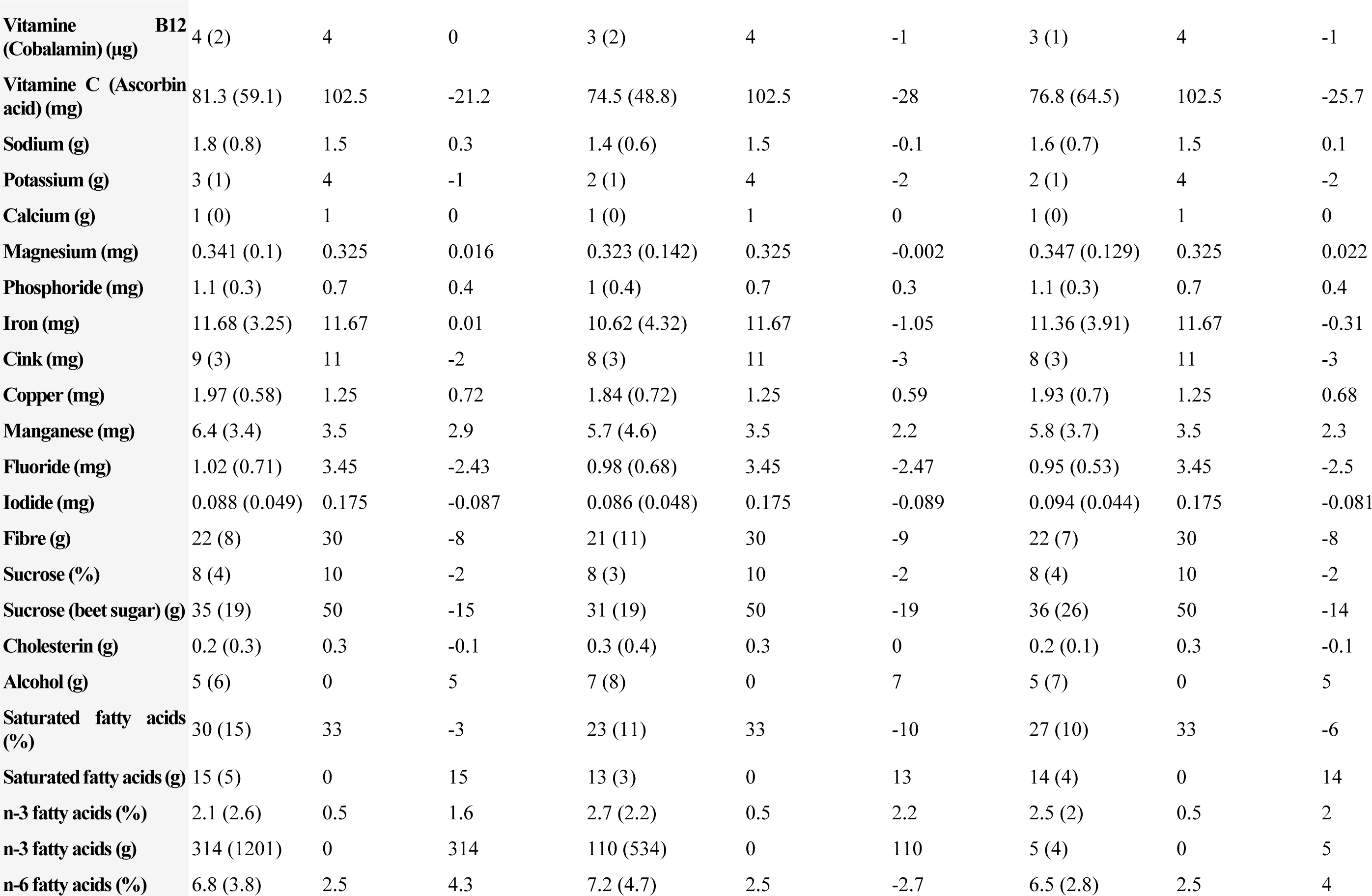
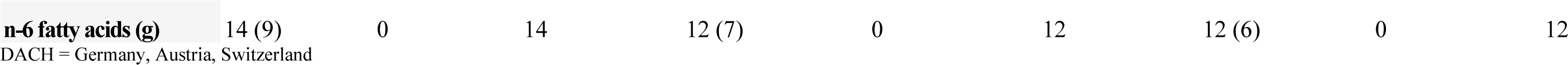
Dietary intake data for fasting diet group.

**Supplemental table 9:**
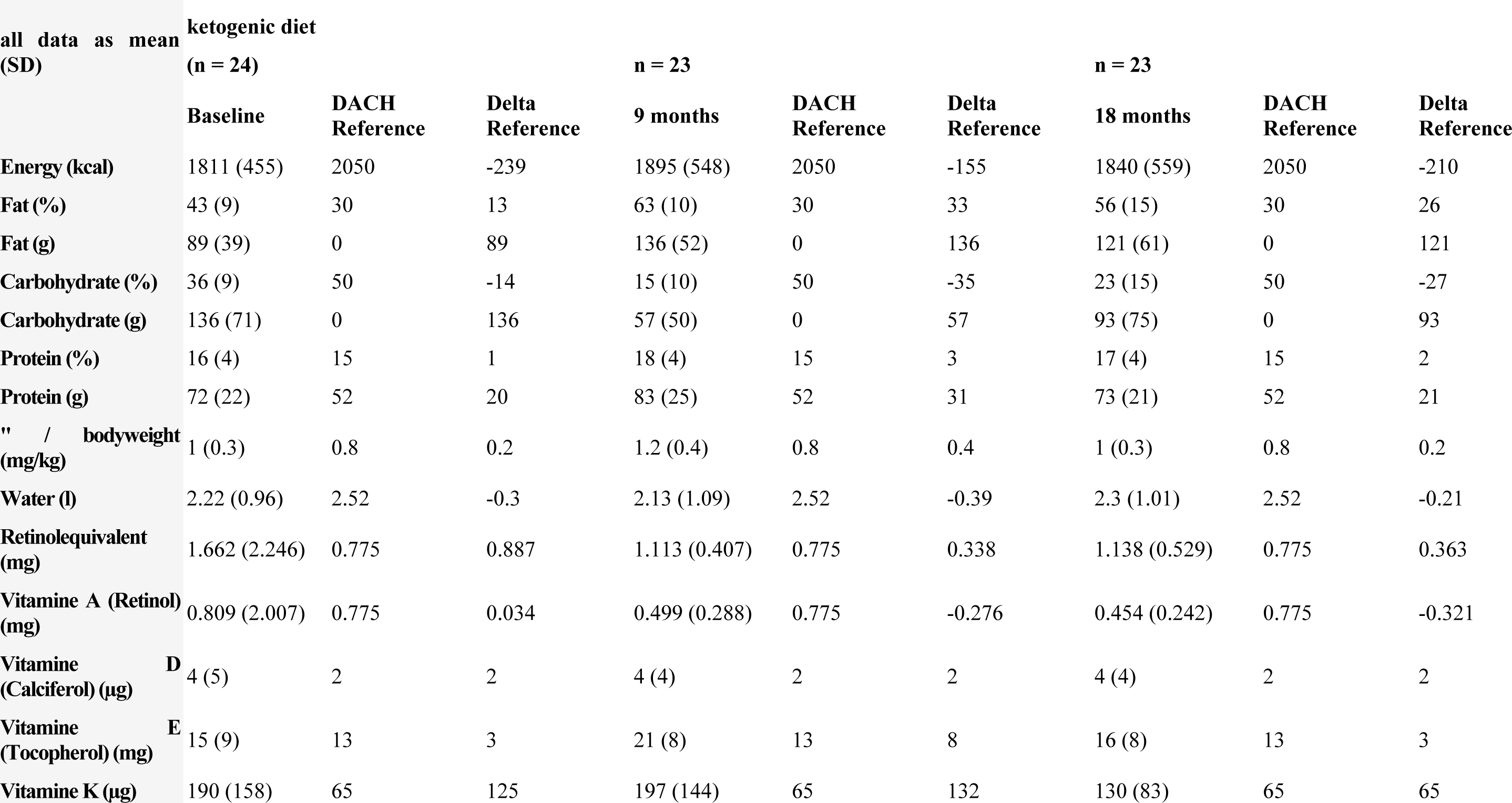
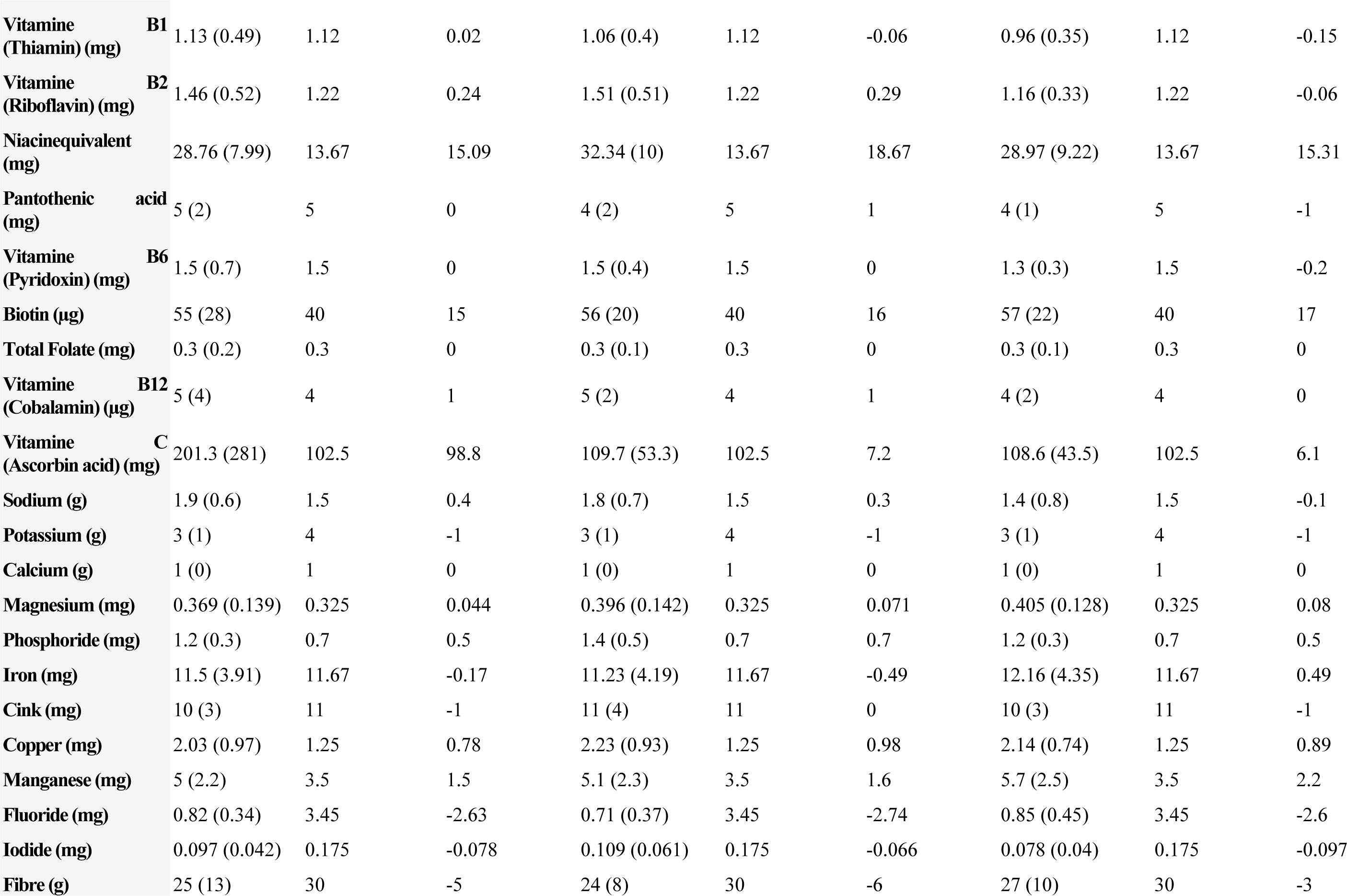
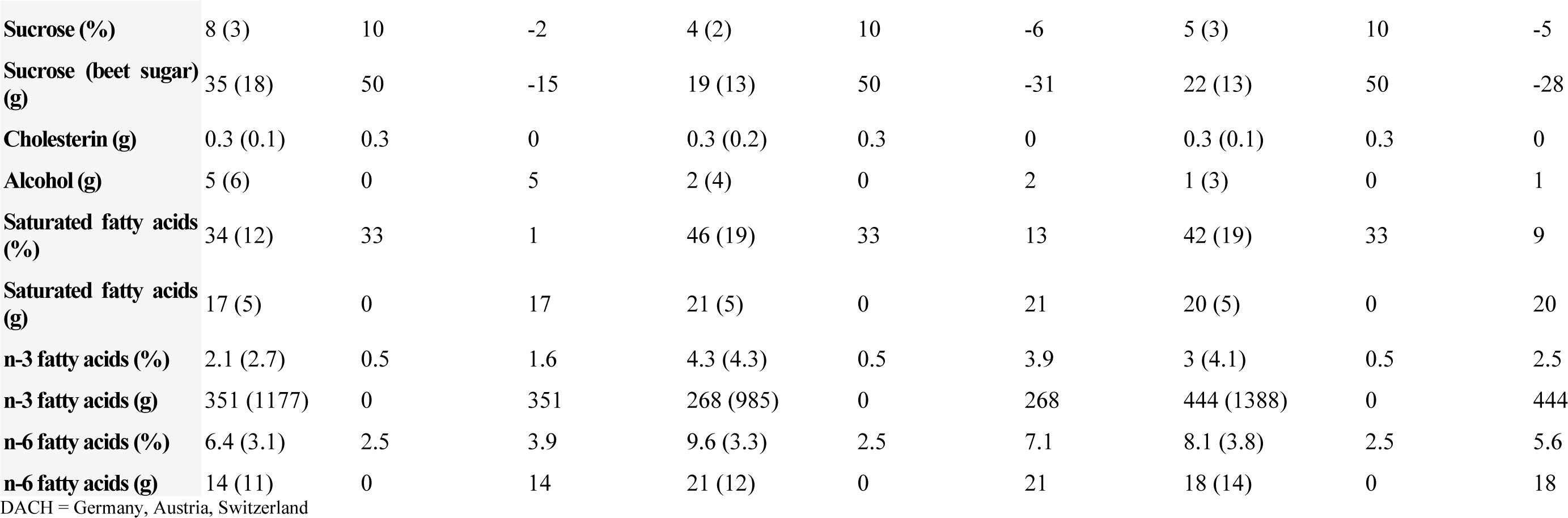
Dietary intake data for ketogenic diet group.

